# IgG4 sensitizes human dorsal root ganglia neurons and patient-derived IgG induces FcγR-dependent pain hypersensitivity in mice

**DOI:** 10.1101/2024.11.12.24317004

**Authors:** Marisol Mancilla Moreno, Cathryn Payne, Brodie J. Woodall, Nathan T. Fiore, Khadijah Mazhar, Alejandro Otero-Pedraza, Asta Arendt-Tranholm, Natalie Yap, Abby P. Chiu, Michael A. Wilde, Pooja J. Patel, Mallory M. Rudd, Diana Tavares-Ferreira, Jeffrey G. Jarvik, Judith A. Turner, Peter M. Grace, George Mount, Anubhav G. Amin, Christoph P. Hofstetter, Theodore J. Price, Muhammad Saad Yousuf, Michele Curatolo

**Affiliations:** Center for Advanced Pain Studies, Department of Neuroscience, University of Texas at Dallas, Richardson, TX, USA; Department of Neurological Surgery, University of Washington, Seattle WA, USA; Department of Anesthesiology and Pain Medicine, University of Washington, Seattle WA, USA; Department of Radiology, University of Washington, Seattle WA, USA; Department of Psychiatry & Behavioral Sciences and Department of Rehabilitation Medicine, University of Washington School of Medicine, Seattle WA, USA; The University of Washington Clinical Learning, Evidence and Research (CLEAR) Center for Musculoskeletal Disorders; Laboratories of Neuroimmunology, Department of Symptom Research, MD Anderson Cancer Center, Houston TX, USA; Division of Rheumatology, University of Washington School of Medicine, Seattle WA, USA

**Keywords:** Immunoglobulin 4, human, pain, dorsal root ganglia, fc gamma receptor

## Abstract

**Background and Objective:** Chronic pain is a common and debilitating condition that affects roughly 25% of the population. Recently, B-cell and immunoglobulin (IgG) signaling have been implicated in preclinical models of neuropathic pain and recent transcriptomic data shows increased number of B-cells and IgGs in the dorsal root ganglia (DRG) of individuals with chronic pain. Here we investigated whether a subclass of IgG, IgG4, sensitizes human DRG neurons and whether IgGs from a patient with elevated expression of *IGHG4* in the DRG induces pain hypersensitivity in mice through Fcγ receptors (FcγR).

**Methods:** We analyzed spatial transcriptomic data from cervical DRGs obtained during arthrodesis for chronic pain, and assessed IgG4 and FcγR1A/CD64 localization by immunohistochemistry. Human organ-donor DRG neurons were treated with recombinant IgG4 containing either a wild-type Fc domain or an aglycosylated Fc domain with reduced FcγR binding and excitability was assessed by whole-cell patch-clamp electrophysiology. Total serum IgG from the patient with high *IGHG4* expression was transferred intrathecally into B-cell-deficient mice or mice lacking active FcγRs after chronic constriction injury.

**Results:** Bilateral C2 DRGs recovered from one of 8 patients with chronic pain had high *IGHG4* expression. *IGHG4*-enriched regions colocalized with a B-cell transcriptomic signature, and IgG4 immunoreactivity was identified in the DRG manifesting as plaque-like structures. Further immunohistochemical analysis confirmed FcγR1A/CD64 to be highly expressed in small-diameter sensory neurons. Wild-type recombinant IgG4 lowered rheobase, increased spontaneous firing, and enhanced current-evoked action potential firing in human DRG neurons compared with vehicle and aglycosylated IgG4. Patient-derived total IgG produced mechanical and thermal hypersensitivity in mature B-cell-deficient mice but not in activate FcγR-deficient mice.

**Discussion:** These findings identify IgG4 enrichment within a human sensory ganglion in chronic pain and demonstrate that IgG4 increases human sensory neuron excitability through an Fcγ receptor-dependent mechanism. Patient IgG also promoted FcγR-dependent pain hypersensitivity *in vivo*, supporting localized humoral immune signaling as a potential mechanism in a subset of patients with chronic pain.

## INTRODUCTION

Chronic pain is highly prevalent and a leading cause of disability^1–3^. Despite its prevalence and burden, the molecular mechanisms that drive human chronic pain remain poorly defined. This mechanistic gap hampers the development of effective pharmacological therapies.

In a recent transcriptomic study of human cervical C2 dorsal root ganglia (DRG), we characterized the cellular and molecular landscape of sensory neurons and non-neuronal populations in patients with acute and chronic neck pain^4^. These datasets allow us to identify differences in gene expression across chronic pain samples that may reveal pain-relevant molecular mechanisms at an individual level. A compelling finding of that study was the association of a B-cell transcriptomic signature with chronic neck pain, suggesting that factors released by B-cells within the DRG may play a key role in sensitizing nociceptors and driving chronic neck pain^4^. Another recent study in rodents^5^ demonstrated that B cells deposit immunoglobulin G (IgG) in the DRGs to cause neuropathic pain via a mechanism that requires FcγR signaling. Importantly, that study also detected an enriched transcriptomic signature of B cells in human DRGs obtained from patients with neuropathic pain due to nerve compression at the thoracic spinal level.

IgG subclasses (1-4) differ substantially in their capacity to engage FcγRs and activate downstream immune pathways^6^. IgG4 is particularly distinct because it weakly activates complement, undergoes Fab-arm exchange, and can engage the high-affinity FcγR1A/CD64 receptor^7^. IgG4 is implicated in IgG4-related disease (IgG4-RD), a systemic fibro-inflammatory condition that is thought to arise from dysregulated immune responses that promote IgG4-producing plasma cells^8^. Whether IgG4 can directly alter the excitability of human sensory neurons through its Fc domain and contribute to pain hypersensitivity remains unknown.

Together, these findings provided the rationale to investigate whether IgG4-FcγR interactions contribute to chronic pain in humans and mice. First, we found high levels of *IGHG4* transcripts and IgG4 immunoreactivity in the C2 DRG of a chronic pain patient undergoing arthrodesis. Second, we confirmed FcγR1A/CD64 protein expression in sensory neurons from human organ donor DRGs. Third, recombinant IgG4 treatment of organ donor-derived DRG neurons caused neuronal sensitization in a Fc-dependent manner. Finally, passive transfer of patient-derived IgG in transgenic mice subject to chronic constriction injury caused mechanical and thermal hypersensitivity through activating FcγRs. This integrated approach has allowed us to identify a previously unrecognized immune-neural interaction that likely underlies chronic pain in a subset of patients susceptible to IgG4 producing B cells.

## METHODS

### Identification and characteristics of patient with high levels of *IGHG4* in the C2 DRG

We identified 1 out of 8 total patients to have high levels of *IGHG4* expression from a previously generated spatial transcriptomics dataset of human C2 dorsal root ganglia^4^ (**Fig. s1-2**). We then looked at the clinical characteristics of this patient.

The study was approved by the University of Washington Internal Review Board (study 10916). The patient with high levels of *IGHG4* in the C2 DRG underwent a posterior C1-T2 fusion and bilateral C1-7 laminectomy, with resection of the C2 DRG bilaterally. He presented severe central stenosis and spinal cord compression (**Fig. s3**). Serum immunoglobulin (Ig) quantification showed elevated Immunoglobulin G subclass 4 levels (89mg/dL; laboratory reference range 4-86 mg/dL) at a follow-up 2.5 years after surgery. Further clinical characteristics are reported in **Supplemental methods** and **Supplemental Tables 1-2.**

### Visium spatial RNA-sequencing analysis

We used the Space Ranger v1.1 outputs previously generated from the Illumina BCL files of the C2 dorsal root ganglia dataset^4^. This pipeline allowed the alignment of the FASTQ files with bright-field microscope images and the human reference transcriptome (GRCh38). Using the Loupe Browser (v8, 10x Genomics), the barcodes were manually classified into single or multiple depending on the number of overlapping neurons. Data were cleaned, normalized and further processed using Python (v3.8 with Anaconda distribution), R (v4.3. or above), and the Seurat (v5) visualization and integration workflow to analyze spatially resolved RNA-seq data. We excluded mitochondrial genes to sufficiently power the statistical analysis of non-mitochondrial genes. Batch effects were corrected using the Harmony R package^9^ (**Fig s3-4**). The Spatially weighted pOissoN-gAmma Regression (SONAR)^10^ model was used for cell-type deconvolution, using a previously published single-cell RNA sequencing dataset from human DRG^5^.

### *IGHG4* expression measurement - Counts per million

To quantify *IGHG4* expression, raw count data for each dorsal root ganglion (DRG) were retrieved using the GetAssayData function in Seurat (v5). For each cell, *IGHG4* expression was normalized to counts per million (CPM). The average CPM value per DRG was calculated by using the following formula:

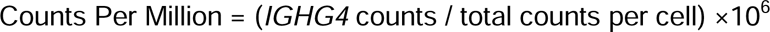

The mean CPM across all cells within the DRG was then computed to provide an averaged *IGHG4* expression level.

### Spatial deconvolution

The spatial deconvolution tool SONAR^10^ has been previously used by our group to deconvolute lumbar DRGs^11^. Now, we used it to estimate the contribution of each cell type to the transcriptomes of each spatial barcode in 3 C2 DRG sections (left and right DRGs from the high-IgG4 donor, and 1 from a control donor). A single-cell RNA-seq dataset of human lumbar DRG^5^ was used as a reference to guide deconvolution, with a curated gene panel for B cell identification (**Supplemental Table 3.**). We generated a raw count matrix with 2993 genes highly enriched per cell type (log2-fold change > 1.0, and adj. p-value < 10-20). All spatial barcodes overlapping tissue were processed with the R script provided by SONAR using default settings, including the pre-processing algorithm that filters out barcodes with low UMI counts.

To infer which cell types are contributing to the high levels of *IGHG4* seen in the C2 DRGs of our patient, we performed linear regression modeling of *IGHG4* expression (in counts per hundred) as a function of the estimated proportion for every cell type. The R lm() function was used to obtain linear regression statistics, including the coefficient, standard error, t-value, p-value, and adjusted r^2^ and the R ggplot package (version 3.5.1) was used to visualize each model. The p-value was used to determine whether the abundance of a cell type correlated with *IGHG4* expression and the adjusted r^2^ was used to determine which cell type was most likely to predict *IGHG4*.

### Organ donor tissue recovery

Lumbar DRGs and lymph nodes were recovered from male and female organ donors (n=7, 42.3 ± 19.9 years old) in collaboration with the Southwest Transplant Alliance and fresh frozen with pulverized dry ice for immunohistochemical experiments or transported in cold artificial cerebrospinal fluid (aCSF) to UTD to proceed with the primary tissue culturing protocol^12^.

### Tissue embedding

Human tissues were embedded in Tissue-Plus O.C.T. Compound (Thermo Fisher Scientific, 23730571) using a cryomold placed over dry ice. The OCT was poured into small volumes surrounding the tissue to avoid thawing. The tissues were sequentially sliced (Leica CM1950) into 20 µm sections at −20°C and mounted onto Superfrost Plus Microscope Slides (Thermo Fisher Scientific, 1255015). High-quality tissue sections containing abundant neurons and no evidence of RNA-compromising artifacts, including cauterization and freezing artifacts, were selected for downstream analysis. Tissue quality and the presence of potential artifacts were assessed by visually examining tissue morphology using a Bio-Rad ZOE Fluorescent Cell Imager. Neurons were identified based on morphology, a cell diameter of approximately 20–110 µm, and the presence of lipofuscin.

### Immunohistochemistry (IHC)

We followed a modified version of the immunohistochemistry protocol described by Yousuf et al.,^13^. Human lymph nodes sections were added as positive control. Primary antibodies (**Sup. Table 4**) were applied overnight at 4°C, followed by incubation with corresponding secondary antibodies for 1 hour at room temperature. Imaging was conducted using an FV4000 confocal laser scanning microscope (Evident Scientific) at 20x magnification. For display purposes, the raw image files were brightened and adjusted for contrast using Olympus CellSens software (v1.18). Refer to **supplementary methods** for further details.

### Primary cultures of human DRG from organ donors and immunoglobulin treatment

Primary cultures were prepared as previously reported^12^. Human DRG cells were incubated with culturing media (1% N2-A, 1% Pen-Strep, 2% SM1, 1% GlutaMAX in BrainPhys Media) for at least 18 hours before being treated. Treatment media was the same as culturing media except that it lacked SM1. Subsequently, human DRG cells were treated overnight (16-18h) between day in vitro 1 and 4 with either 1X sterile Dulbecco’s Phosphate Buffered Saline (Corning, 21-031-CV) as a vehicle, 10 nM human Control Antibody IgG4 Wildtype Fc (AB biosciences, PZHU003, rIgG4-WT), or 10 nM of human Control Antibody IgG4 Aglycosylated Fc (AB biosciences, PZHU004, rIgG4-mut).

### Organ donors: Patch clamp electrophysiology

Whole-cell patch clamp electrophysiology was performed as described previously^14^. Human DRG neurons were plated at 50 neurons per coverslip on 18 mm coverslips. Recordings were obtained using a Multiclamp 700B amplifier in conjunction with pClamp10 acquisition software. Glass micropipettes (Sutter instrument company, BF150-110-10) were used to apply suction until a giga-ohm seal was attained and the membrane was ruptured to achieve whole-cell configuration. Cell health was assessed in voltage clamp using a −70 mV to 0 mV to −70 mV voltage step to test for presence of voltage-driven inward and outward currents. Refer to **supplementary methods** file for further details on these experiments and **Sup. Table 5** for donor’s demographic information.

### Animals

Male and female mice lacking mature B cells (muMT mice; Jackson Laboratory, strain B6.129S2-*Ighm^tm1Cgn^*/J, #002288)^15^, or mice lacking activating Fcγ receptors (FcγR^-^ mice; Taconic Biosciences, strain B6.129P2-*Fcer1g^tm1Rav^*N12, #583)^16^ were used for all experiments. Mice were bred in-house at the University of Texas MD Anderson Cancer Center Association for Assessment and Accreditation of Laboratory Animal Care-accredited vivarium. Animals were housed in the specific pathogen-free MD Anderson vivarium with *ad libitum* access to food and drinking water on a 12 h light-dark cycle. Mice were at least 8 weeks old at the start of experimental manipulations. Animals were randomly assigned to treatment groups using an online group randomization tool (randomlists.com/team-generator). All animal experiments and procedures were approved by the Institutional Animal Care and Use Committee of the University of Texas MD Anderson Cancer Center.

### Surgery in the mouse model

Neuropathic pain was modelled in mice (n=17) using unilateral sciatic nerve chronic constriction injury (CCI) as previously described^5^. Mice were anesthetized with inhaled isoflurane (2.5% in 2 L/min oxygen for induction and maintenance) and placed on an electric heating pad to maintain body temperature. The left hindlimb was shaved and disinfected with povidone-iodine followed by 70% ethanol. An incision was made to expose the left sciatic nerve and polypropylene pipette tips were used to bluntly isolate the nerve proximal to the trifurcation. Three ligatures (5-0 chromic gut; Ethicon) were loosely tied around the sciatic nerve approximately 1 mm apart. The muscle layer was sutured closed (4-0 Vicryl; Ethicon), and the skin was closed with a 9-mm wound clip. At the completion of surgery, mice were returned to their home cages without post-surgical analgesic treatment and were monitored postoperatively until fully ambulatory. Mechanical allodynia was assessed using von Frey filaments with the up-down method as previously described^5^. Briefly, 8 von Frey filaments ranging from 0.02 g to 1.4 g were applied, beginning with the 0.16 g filament, and 50% withdrawal thresholds were calculated. Paw withdrawal latency to radiant heat was measured using the Hargreaves device (IITC Life Science Inc.), as previously described^5^.

### IgG purification

Blood was centrifuged at 3214 x g at 4°C for 10 minutes, and serum was collected and stored at −80 °C. Patient IgG was purified from serum using HiTrap MabSelect SuRe 70 1 mL columns (Cytiva 11-0034-93) according to the manufacturer’s instructions. Serum (2 mL) was diluted 1:5 in sterile 1x PBS (pH 7.4) and loaded onto the HiTrap column. The column was washed with 5 mL of 20 mM sodium phosphate and 0.15 M sodium chloride, followed by 3 mL of 50 mM sodium acetate (pH 6). IgG was eluted with 5 mL sodium acetate (pH 3.5) and 5 mL IgG elution buffer (pH 2.8; ThermoFisher Scientific, 21004). Approximately 10-12 fractions (1 mL each) were collected in tubes containing 100 µL of 1 M Tris-HCl (pH 9) to neutralize the eluate. Fractions were pooled and dialyzed overnight at 4°C against 1x PBS (pH 7.4) using Slide-A-Lyzer dialysis cassettes (2kDa MWCO; ThermoFisher Scientific, 66203). Protein concentration was measured at 280 nm using a NanoDrop spectrophotometer (Thermo Fisher Scientific). IgG samples were concentrated to 10 µg/µL using Amicon Ultra-4 10kDa centrifugal filters (Millipore Sigma, UFC801008), sterilized by filtration through 0.2 µm filter, and stored at −80°C until use.

### Passive transfer of IgG

IgG was delivered to mice intrathecally (i.t.) as previously described^5^. Mice were anesthetized with inhaled isoflurane (2-3% in oxygen for induction and maintenance) and placed on an electric heating pad to maintain body temperature. Fur along the lumbar vertebrae was shaved with an electric razor and the skin was cleansed with 70% ethanol. Custom injectors were constructed by fitting a 27-gauge needle into polyethylene tubing (PE-20) and sterilized by soaking in 70% ethanol followed by overnight drying. IgG was drawn into the injector using a 25 µL Hamilton syringe and the injector was inserted into the intrathecal space above the lumbar enlargement. Patient IgG or human IgG4 isotype control (Bio X Cell, #CP147) (40 µg in 4 µL) was slowly injected over approximately 2 minutes, after which mice were returned to their home cages. Each mouse received daily injections for three consecutive days (days 14-16 after CCI). Baseline behavioral measurements were obtained on day 13 after CCI (24 hours before the first injection), and post-treatment behavior was assessed on day 17 after CCI 24 hours after the final injection.

### Behavioral tests for mechanical allodynia and thermal hyperalgesia

Animals were habituated to the testing apparatus and experimenter prior to testing. The experimenter was blinded to group assignment throughout behavioral assessments. Mechanical allodynia was assessed using von Frey filaments with the up-down method as previously described^5^. Briefly, eight von Frey filaments ranging from 0.02 g to 1.4 g were applied, beginning with the 0.16 g filament, and 50% withdrawal thresholds were calculated.

Paw withdrawal latency to radiant heat was measured using the Hargreaves device (IITC Life Science Inc.) as previously described^5^. Mice were placed on a pre-warmed glass plate (29-31 °C) within transparent enclosures. A halogen light beam (set to an intensity of 20% of maximum) was applied to the plantar surface of both hind paws. A cutoff time of 20 seconds was used to prevent potential overheating and tissue damage. Average withdrawal latencies for each mouse were calculated from three trials separated by intervals of 5-10 minutes.

### Statistical analysis

Statistical analyses were performed using GraphPad Prism. For electrophysiological experiments rheobase comparisons, an ordinary one-way ANOVA followed by Tukey’s multiple comparisons test was used to assess differences between groups. The proportion of neurons exhibiting spontaneous action potential firing was compared descriptively across groups. For current ramp–evoked firing, a two-way ANOVA followed by Tukey’s multiple comparisons test was used. Statistical significance was defined as p < 0.05. Group sizes were as follows: vehicle (n = 28 for rheobase and spontaneous firing; n = 26 for current ramps), rIgG4-mut (n = 27 for rheobase, spontaneous firing and current ramps), and rIgG4-WT (n = 26 for rheobase and spontaneous firing; n = 22 for current ramps).

Statistical analyses for mechanical allodynia (von Frey, 50% withdrawal threshold in grams) and thermal hyperalgesia (Hargreaves, withdrawal latency in seconds) were analyzed using two-way ANOVA followed by Sidak’s multiple comparisons test to assess treatment effects over time. In addition, pre- and post-treatment responses (24 h after the third intrathecal injection) were compared within groups using a paired two-tailed Student’s t-test with Welch’s correction. Statistical significance was defined as p < 0.05. Sample sizes were n = 5–6 per group across conditions.

## RESULTS

### Spatial transcriptomics revealed high levels of *IGHG4* in a DRG from cervical arthrodesis patient

Bilateral DRGs were resected from patients (n=8) undergoing arthrodesis surgery due to their chronic neck pain. We previously performed spatial transcriptomics on these DRGs^4^ and noted that DRGs from one patient had unusually high levels of *IGHG4* in the C2 DRGs (**Fig s1-2**). Expression levels varied significantly between the two sides of this patient’s neck, with 23,775 counts per million (CPM) on the right side compared to 1,720 CPM on the left (**Fig. 1a-b and Fig. s4-5**).

**Figure 1.**
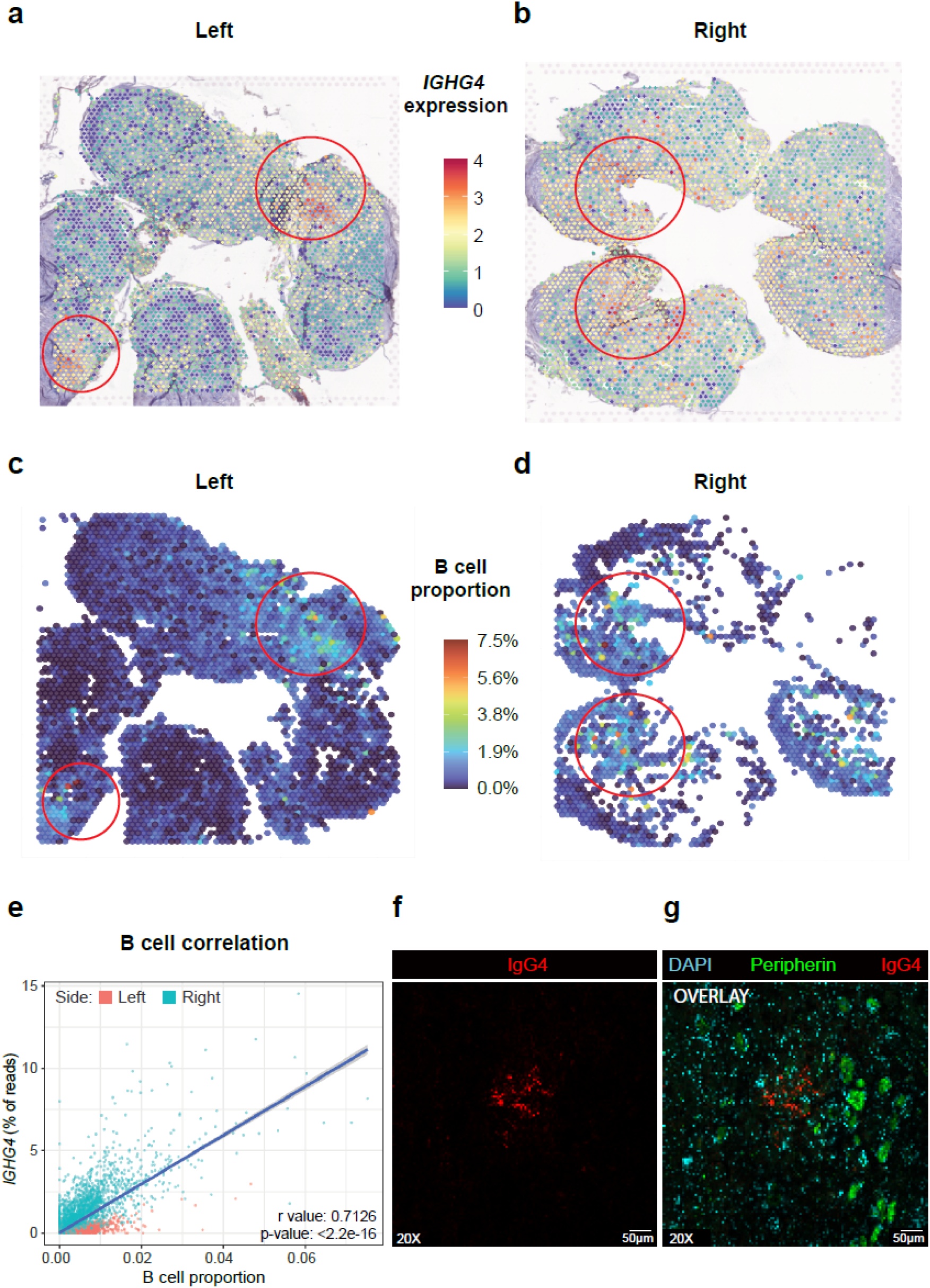
Spatial sequencing identification of *IGHG4*, computational cell type deconvolution, and validation at the protein level with immunohistochemistry. **a and b** DRG sections mounted on 10X Genomics Visium slides with overlapping barcodes. *IGHG4* expression is shown on a scale from 0 (blue) to 4 (red). Red circles indicate areas of high *IGHG4* expression. **c and d** DRG computational cell type deconvolution showing an increased proportion of B cells in areas indicated with red circles. The scale ranges from 0% (dark blue) to 7.5% (dark red). **e** Correlation plot between B cell proportion and the percentage of *IGHG4* transcript reads on both sides. R=0.7126, p<2.2e-16. **f** Validation of *IGHG4* at the protein level. Immunofluorescence of IgG4; **g** overlay of DAPI (cyan) to identify nuclei, Peripherin (green) as a neural marker, and IgG4 (red).

To investigate the source of *IGHG4* expression, we performed spatial deconvolution. Regions with high *IGHG4* expression were correlated significantly with B cell abundance (r = 0.71 for the right side and r = 0.60 for the left side, both p-values < 0.05); **Fig.1c-e**), but not with any other cell type (**Fig.s6-8**). This indicates that *IGHG4* expression was likely derived from B cells, which are crucially involved in IgG4 production^5^.

To examine protein expression, we used an IgG4-antibody validated through positive staining in human lymph node (**Fig. s9**). In the right DRG we observed IgG4 immunoreactivity in a plaque-like structure (**Fig.1f and g**). These observations laid the foundation of our hypothesis that IgG4 may sensitize DRG neurons and contribute to pain hypersensitivity.

### Immunohistochemical analysis confirms FcγR1A/CD64 expression in human DRG neurons from organ donors

To determine whether the high-affinity IgG receptor, FcγR1A/CD64, is expressed by neurons, we evaluated FcγR1A/CD64 expression using immunohistochemistry of human DRG sections from organ donors (n=2). FcγR1A/CD64 was detected predominantly in neurons with prominent membranous staining and diffuse cytoplasmic staining (**Fig. 2a**). We also noted a higher expression of FcγR1A/CD64 in small-diameter sensory neurons and putative nociceptors (20-50 µm) (**Fig. 2b**). This pattern was consistent previously published spatial sequencing data from human organ donor DRGs which showed that *FCGR1A* transcripts were found in multiple classes of nociceptors^17^ (**Fig. 2c**). These findings demonstrate that FcγR1A/CD64 is expressed by human sensory neurons and support the potential for direct IgG4–FcγR1A/CD64 interactions to modulate neuronal function.

**Figure 2.**
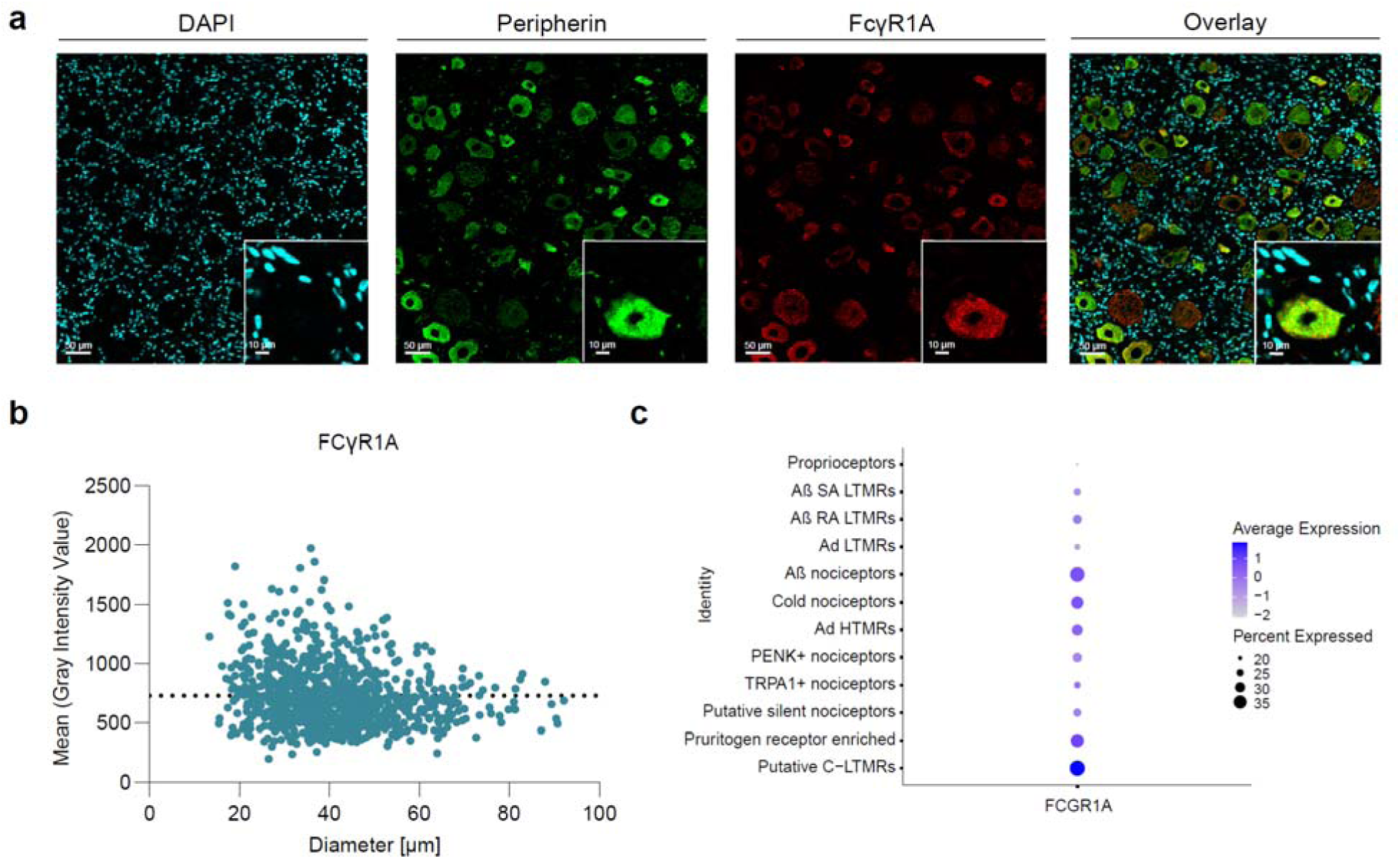
Immunohistochemical analysis confirms FcγR1A/C64 expression in human DRG neurons from organ donors. **a** Representative immunofluorescence images of DRG sections stained with DAPI (cyan, nuclei), peripherin (green, neuronal marker) and FcyR1a (red), demonstrating FcγR1A/CD64 expression throughout the cell bodies of sensory neurons. **b** Quantification of FcγR1A/CD64 expression across neuronal sizes revealed a higher expression in small-diameter neurons (20-50 µm) (Data presented as mean ± SEM; n=2 donors; dotted line represents average), consistent with **c** Visium spatial sequencing data from human organ donor DRGs^17^.

### Whole-cell patch-clamp electrophysiology demonstrates IgG4-induced hyperexcitability in human sensory neurons

To test whether monomeric IgG4 binding to FcγR is sufficient to induce neuronal hyperexcitability, we treated cultured human sensory DRG neurons (5 donors, 50 cells per donor) with recombinant human IgG4 antibodies engineered with silenced Complementary Determining Regions (CDRs) and either a wild-type Fc domain ((rIgG4-WT; AB Biosciences, PZHU003) or an aglycosylated Fc domain with the N297A mutation (rIgG4-mut; AB Biosciences, PZHU004), along with sterile DPBS as a vehicle control, and subjected them to whole-cell patch-clamp electrophysiology. Because these antibodies have silenced-CDRs, they cannot bind antigen or form immune complexes, allowing us to interrogate the effect of monomeric IgG4 binding to FcγRs. Binding properties reported by the manufacturer (AB biosciences) indicate that rIgG4- WT binds FcγRI with high affinity and other FcγRs with low affinity, whereas rIgG4-mut, which has an aglycosylated Fc domain, binds poorly to all FcγRs. Human DRG neurons treated with rIgG4-WT had a lower rheobase **(Fig. 3a)**, showed a greater proportion of spontaneous firing **(Fig. 3b and Fig. s10)** and fired more action potentials in response to current ramps **(Fig. 3c)** compared to both vehicle and rIgG4-mut treatments. Other potential action parameters such as amplitude and half-width were not altered significantly **(Table 1.).**

**Figure 3.**
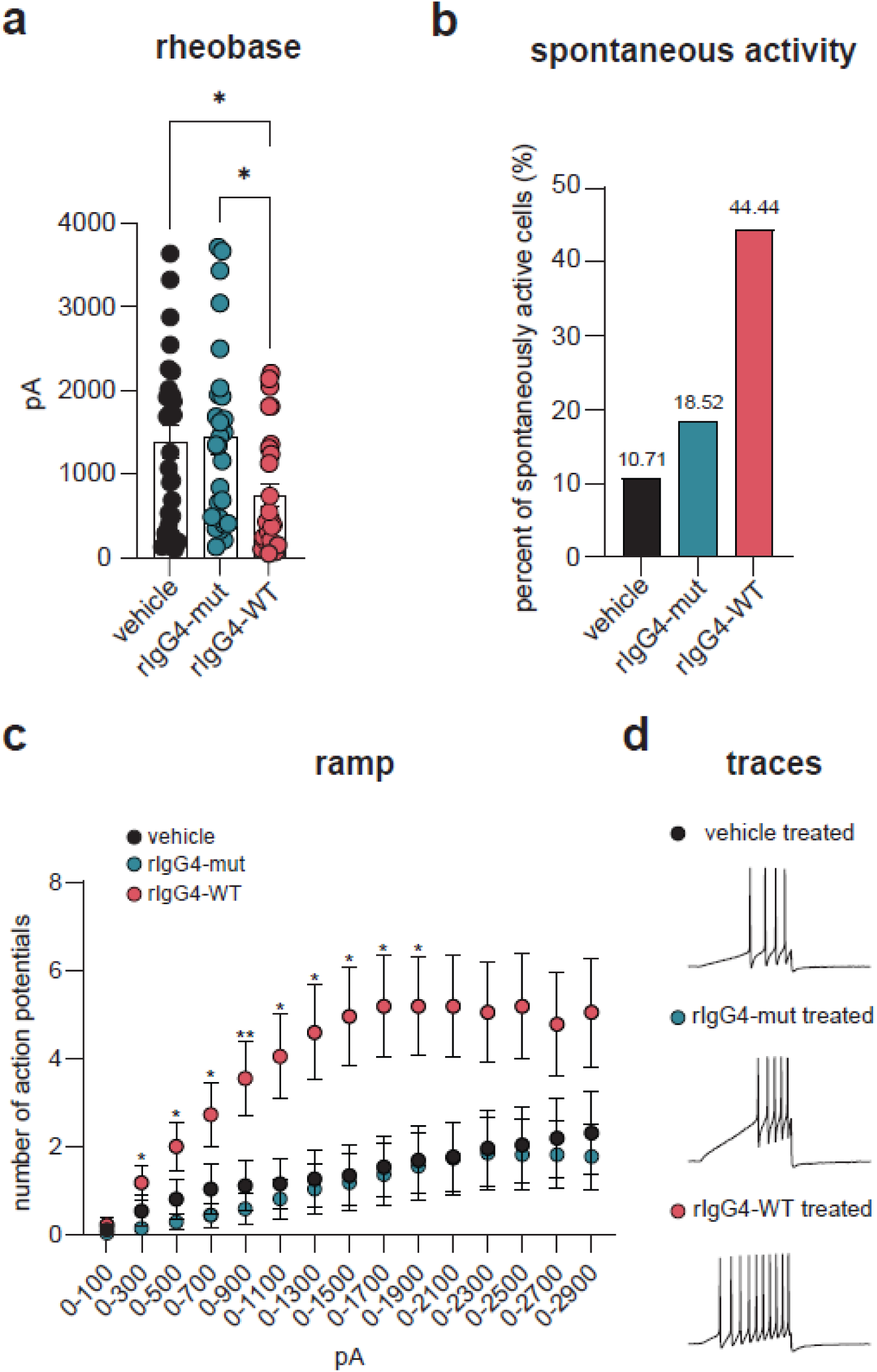
rIgG4 with wild-type Fc domain induces hyperexcitability in human sensory neurons. **a** rIgG4-WT treatment significantly reduced rheobase compared with vehicle-treated neurons. Statistical analysis was performed using ordinary one-way ANOVA followed by Tukey’s multiple comparisons test (vehicle vs. rIgG4-WT, p = 0.0370, rIgG4-WT vs rIgG4-mut, p = 0.0222). Group sizes: vehicle n=28; rIgG4-mut n=27; rIgG4-WT n=26. **b** A greater proportion of rIgG4-WT treated neurons exhibited spontaneous action potential firing relative to vehicle and rIgG4-mut controls. Group sizes: vehicle n=28; rIgG4-mut n=27; rIgG4-WT n=26. **c** DRG neurons treated overnight with rIgG4-WT fired significantly more action potentials in response to depolarizing current ramps as compared with vehicle and rIgG4-mut treated controls. Statistical analysis was performed using 2-way ANOVA followed by Tukey’s multiple comparisons (*= p<0.05, **=p<0.005). Group sizes: vehicle 26, rIgG4-mut 27, rIgG4-WT 22. **d** Representative traces from current ramps illustrating firing patterns of neurons per group.

**Table 1.**
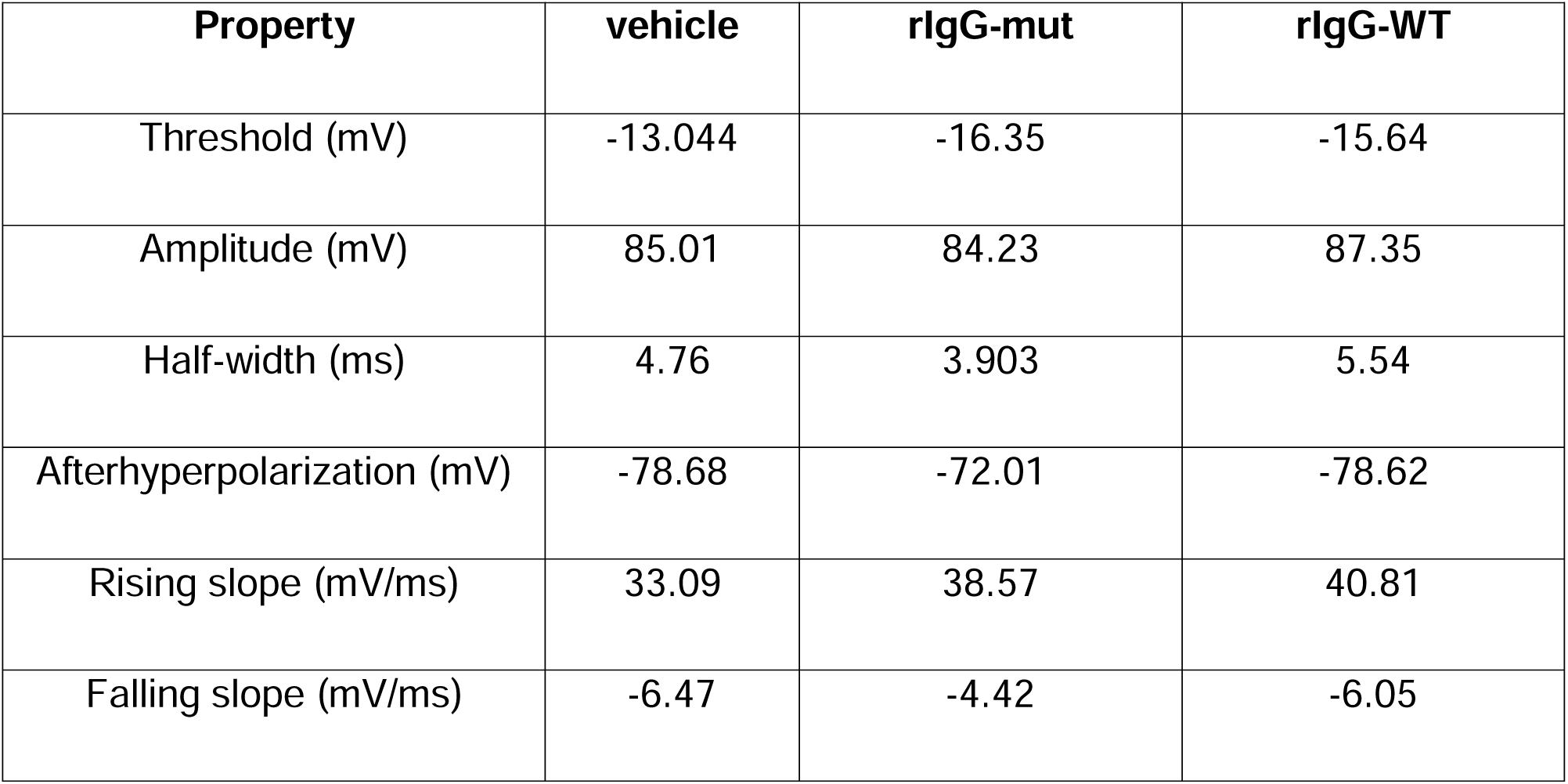
Additional parameters of electrophysiological recordings.

| Property | vehicle | rlgG-mut | rlgG-WT |
| --- | --- | --- | --- |
| Threshold (mV) | -13.044 | -16.35 | -15.64 |
| Amplitude (mV) | 85.01 | 84.23 | 87.35 |
| Half-width (ms) | 4.76 | 3.903 | 5.54 |
| Afterhyperpolarization (mV) | -78.68 | -72.01 | -78.62 |
| Rising slope (mV/ms) | 33.09 | 38.57 | 40.81 |
| Falling slope (mV/ms) | -6.47 | -4.42 | -6.05 |

### Patient-derived IgG passive-transfer behavioral assessment reveals FcγR- dependent mechanical allodynia and thermal hyperalgesia in the mouse CCI model

To address whether patient-derived IgG were sufficient to cause mechanical allodynia and thermal hyperalgesia, we performed a passive intrathecal transfer of purified total IgG isolated from patient serum into mice subjected to the chronic constriction injury (CCI) pain model. In CCI, nerve injury induces local B-cell activation and endogenous IgG production, which generates pain-like behaviors at the time of injury^5^. Importantly, our previous studies showed that intrathecal transfer of IgG isolated from CCI mice induces mechanical allodynia in nerve-injured, but not naïve mice. These findings indicate that the pronociceptive actions of IgG require an injury-associated environment, potentially involving recognition of injury-associated antigens and/or increased availability of activating FcγRs. To isolate the effects of exogenous IgG in this context, we performed CCI on two genetically modified mouse strains: muMT mice, which lack mature B cells and therefore do not produce endogenous IgG, and FcγR^-^ mice, which lack activating Fcγ receptors. Since either the IgG antibody producing B-cells (muMT) or the IgG receptor (FcγR^-^ mice) are lacking, mice with these mutations do not develop mechanical and thermal hypersensitivity. We thus hypothesized that exogenous administration of patient-derived IgG in muMT mice, but not FcγR^-^ mice, would restore pain-like behaviors.

We used human IgG4 S228P as an isotype control because it has minimal antigen binding since the S228P mutation prevents Fab-arm exchange. In von Frey testing, CCI muMT mice that received patient-derived IgG developed significant mechanical allodynia on days 15-17. In contrast, CCI FcγR^-^ mice receiving the same patient-derived IgG did not show significant changes in withdrawal thresholds compared to isotype-treated controls or muMT mice **(Fig. 4a and 4b)**. Mice receiving patient-derived IgG also exhibited significant thermal hyperalgesia in the Hargreaves assay 24 hours after the final injection relative to pre-injection withdrawal latency **(Fig. 4c and 4d)**. These data suggest that patient-derived IgG is sufficient to cause pain hypersensitivity in mice and that FcγRs are necessary for this effect.

**Figure 4.**
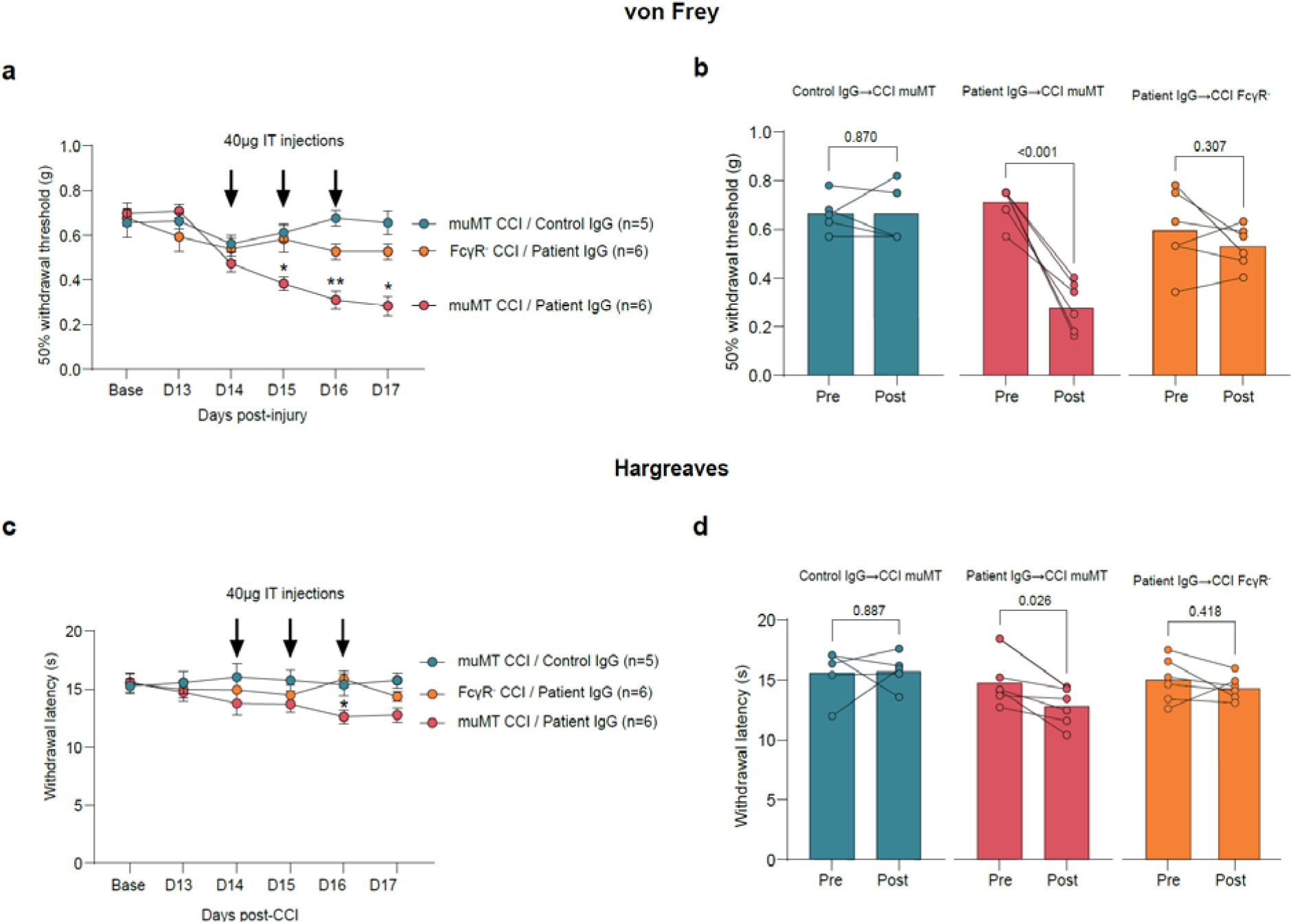
Passive transfer of patient IgG induces mechanical allodynia and thermal hyperalgesia in mice lacking B cells (muMT) subjected to the chronic constriction injury (CCI) pain model. Patient-derived or control IgG was passively transferred to CCI muMT or FcγR^-^ recipient mice (three daily 40 µg intrathecal injections starting 14 days after CCI). **a** von Frey thresholds for mechanical allodynia were measured in ipsilateral hindpaws. Arrows indicate daily IgG injection timepoints. **b** 50% withdrawal thresholds measured by von Frey testing in the ipsilateral hindpaw before (pre) and 24 hours after (post) the third IgG injection. Patient IgG transfer significantly reduced withdrawal thresholds in CCI muMT mice. **c** Hargreaves paw withdrawal latencies for thermal hyperalgesia were measured in ipsilateral hindpaws. Arrows indicate injection timepoints. **d** Hargreaves paw withdrawal latencies before (pre) and 24 hours after (post) the third daily injection of IgG. Patient IgG transfer significantly reduced withdrawal latencies in CCI muMT mice. Statistical analysis was performed using (**a and c**) 2way ANOVA followed by Sidak’s multiple comparisons test (muMT CCI / Patient IgG vs muMT CCI / Control IgG *= p<0.05, **=p<0.005) or (**b and d)** paired two-tailed Student’s *t*-test (pre vs post IgG injection). *n* = 5-6 per group.

## DISCUSSION

B cells and their immunoglobulin G products (IgG) have been shown to accumulate in sensory tissues and contribute to the development of pain-like behaviors in mice through IgG-FcγR signaling in lumbar DRG and spinal cord^5,18^. This mechanism has been demonstrated across multiple murine pain models, including sciatic nerve chronic constriction injury (CCI), where nerve injury leads to local B-cell-mediated production of IgG, subsequent FcγR engagement, and pain hypersensitivity^5^. Analysis of bulk RNA- sequencing data from thoracic vertebrectomy patients with and without neuropathic pain suggests that infiltration, activation and differentiation of B cells into plasmablasts in the human DRG is associated with neuropathic pain in this patient population^5^.

IgG subclasses vary in their effector functions, receptor affinities, and inflammatory potential, suggesting that individual subclasses may differentially influence neuronal activity and shape distinct pain phenotypes^6^. IgG4 is a subclass of IgG present in humans but not in mice, characterized by its limited capacity to activate complement C1q and its ability to undergo Fab-arm exchange, a process that generates functionally monovalent, bispecific antibodies^7^. Immunoglobulins are known to form immune- complexes in the presence of an antigen but unlike IgG1 and IgG3, IgG4 can also form Fc-mediated aggregates through extensive Fc-Fc interactions, providing a potential mechanism for pathogenic activity even in the absence of antigen^7^.

Recent data show a critical role of B cells in promoting neuropathic pain via IgG accumulation in the mouse and human DRG^5^ and we found co-localization of B cells and *IGHG4* expression in the DRGs of our patient. In another recent study that collected sera from patients with and without neuropathic pain for in-vitro analyses, anti-contactin- associated protein-like 2 (CASPR2) IgG4 increased DRG neuronal excitability by modifying potassium channel function^19^. These data provide support for a role of B cell infiltration and differentiation and *IGHG4* expression in pain.

Our electrophysiology data demonstrate that rIgG4-WT is sufficient to increase human sensory neuron excitability, indicating that neither antigen engagement nor immune complex formation is required. While untested in this paper, one possible explanation for this observation is that these recombinant IgG4 molecules create self-aggregates sufficient to engage FcγR signaling and subsequent neuronal hyperexcitability.

Complementing these findings, *in vivo* passive transfer of patient-derived IgG induced robust mechanical allodynia and thermal hyperalgesia in muMT mice following CCI, but not in FcγR- mice. This demonstrates that Fcγ receptor activation is required to elicit pain-like behavior, as previously shown in similar studies exploring mouse DRG at the lumbar level^5^.

A relevant question is whether biomarkers, such as serum IgG4 levels, can be used to identify IgG4 involvement in patients with chronic pain. The availability of such biomarkers could enable more targeted and mechanism-based therapeutic strategies that are not currently part of standard treatment paradigms for chronic neck pain, including B cell depletion by inebilizumab^20^, intravenous immunoglobulin^21^, and other emerging treatments of IgG4-RD^22,23^.

A major strength of this work is the integrated approach combining spatial transcriptomics of DRG from rigorously-phenotyped patients with chronic pain, immunohistochemistry of human DRG sections, patch-clamp electrophysiology in human organ donor DRG neurons, and transfer experiments in a mouse model of neuropathic pain. Together, these complementary methodologies provide strong mechanistic support for a pathogenic role for IgG4-FcγR interactions in chronic pain.

This study has some limitations. First, we could analyze only the C2 DRG and thus cannot confirm *IGHG4* expression in DRGs in other spinal levels or in other tissues. Infiltration of IgG4-positive plasma cells in sural nerves has been reported in patients with sensory-motor impairments^8^ and it is conceivable that *IGHG4* expression in the DRGs is ubiquitous, contributing to the patient’s pain and neurologic symptoms involving all 4 extremities. In addition, we did not measure the local concentration of IgG4 within the DRG. Since the DRG has a relatively permeable blood-DRG barrier that permits circulating immunoglobulins greater access than the blood-brain barrier, we postulated that increases in serum IgG4 levels would reflect changes in tissue concentrations or local IgG4 accumulation. Second, our electrophysiology data with IgG4 and profuse immunostaining of FcγR1A/CD64 in sensory neurons (**Fig. 2**) suggests that a potential mechanism of sensitization of human sensory neurons is a direct IgG4-mediated activation of FcγRs expressed in sensory neurons. However, we cannot rule out an indirect mechanism where non-neuronal cells *in vitro* may be stimulated with IgG4 and subsequently release pro-inflammatory factors underlying neuronal hyperexcitability. Third, for the passive-transfer experiments, we used patient-derived total IgG rather than purified IgG4. The limited volume of available patient serum, together with the relatively low abundance of IgG4 within the total IgG pool, precluded recovery of sufficient purified IgG4 for the in vivo experiments. Additionally, Protein A-based purification method use here is known to efficiently enrich IgG1, IgG2, and IgG4, the subclass of primary interest here, while IgG3 recovery is comparatively lower. Thus, the subclass composition of the injected preparation may not completely reflect the patient’s repertoire, with IgG3 potentially underrepresented. Nonetheless, the predominance of *IGHG4* transcripts in the sequencing data and elevated blood levels of IgG4 indicate that IgG4 is likely a major contributor to the observed phenotype.

We demonstrated that recombinant wild-type IgG4 increases neuronal excitability in vitro, while IgG from a patient with markedly elevated *IGHG4* expression in a human cervical DRG induced mechanical allodynia and thermal hyperalgesia in mice following chronic constriction injury through FcγR-dependent signaling. Together, these findings support a pathogenic role for IgG4-FcγR interactions in chronic pain and associated neurological symptoms and suggest new opportunities for improved diagnosis and mechanistically targeted therapies in patients with chronic pain who are susceptible to B-cell infiltration, activation and differentiation into plasma cells, along with increased *IGHG4* expression.

## DATA AVAILABILITY

Processed data are publicly accessible on the SPARC portal; DOI:10.26275/mfxc-k28b.

## ACKNOWLEDGEMENTS

We thank the patients and the clinical staff who made this research possible.

## FUNDING

NIH: R01AR078192 (MC, TJP, CPH); NIH: U19NS130608 (MC, TJP, CPH); NIH - P30 AR072572-06 (JGJ, JAT, MC); NIH: R01NS126252 (PMG); Department of Defense HT9425-24-1-0109 (PMG); NIH: R01DK134893 (MSY).

## COMPETING INTERESTS

Theodore J. Price is a co-founder of and holds equity in NuvoNuro, PARMedics, and Nerveli and has received research grants from AbbVie, Eli Lilly, Grünenthal, Evommune, Hoba Therapeutics, and The National Institutes of Health; Muhammad Saad Yousuf is a co-founder of and holds equity in NuvoNuro. All other authors have no disclosures.

## Supplementary Figures

**Figure s1.**
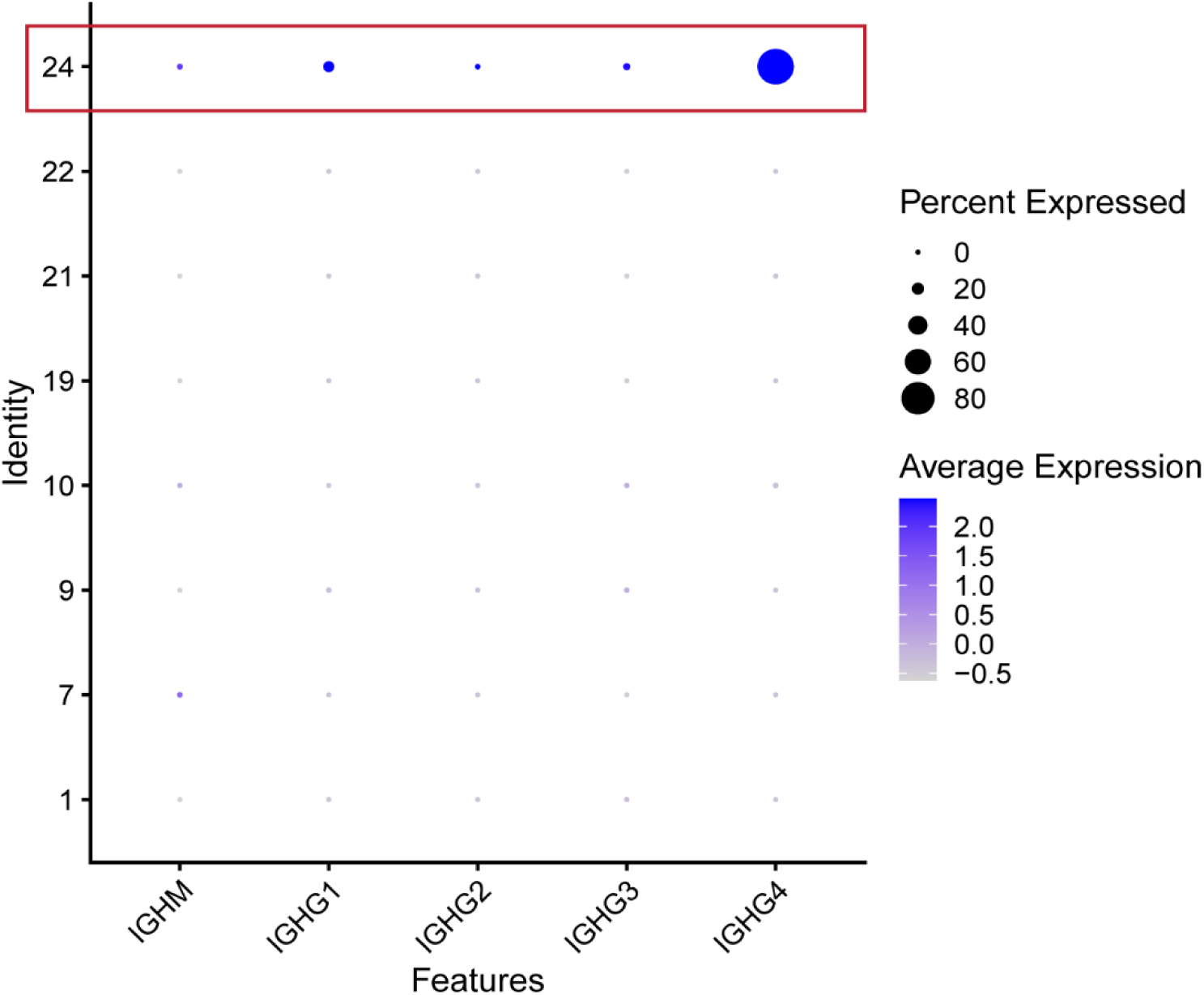
Comparison of selected immunoglobulin expression across patients DRGs. Dot plot showing the transcriptomic expression of selected immunoglobulin transcripts. Average expression indicated by a gray-blue gradient, with darker blue representing higher expression. Circle size is proportional to the percent of barcodes expressing the indicated transcript. Patient DRGs are numbered according to the patient labelling used by Arendt-Tranholm, et. al (2025). Patient 24 corresponds to the case study patient with elevated *IGHG4* levels.

**Figure s2.**
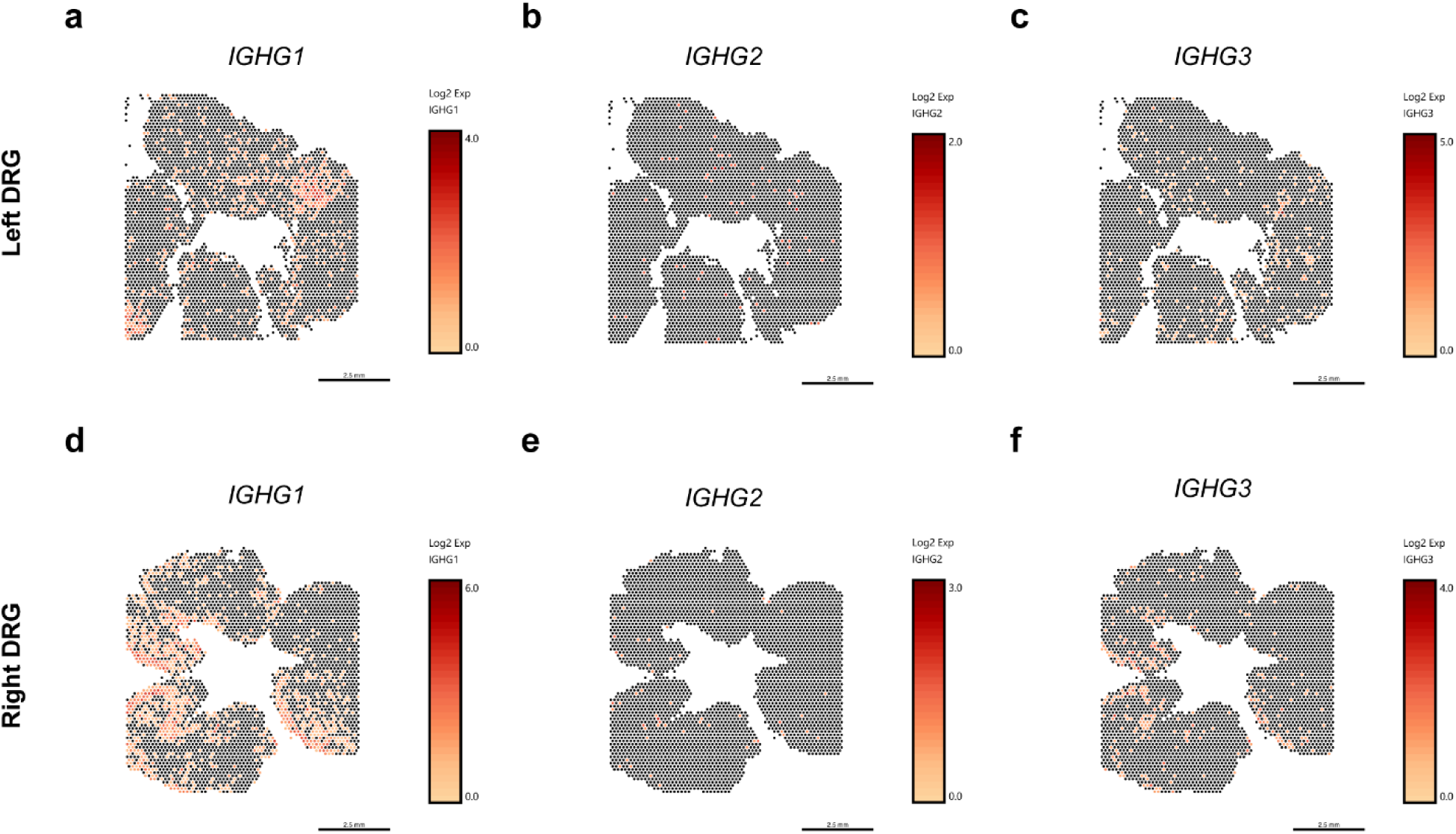
Localized gene expression of select immunoglobulins in right and left DRG of case study patient. **a-c** Spatial transcriptomic localization and log2- transformed expression of *IGHG1*, *IGHG2* and *IGHG3,* respectively. Barcodes with higher expression are shown on a yellow-to-red gradient, with red indicating the highest expression. **d-f** Corresponding panels for the right DRG. Scale bar = 2.5 mm.

**Figure s3.**
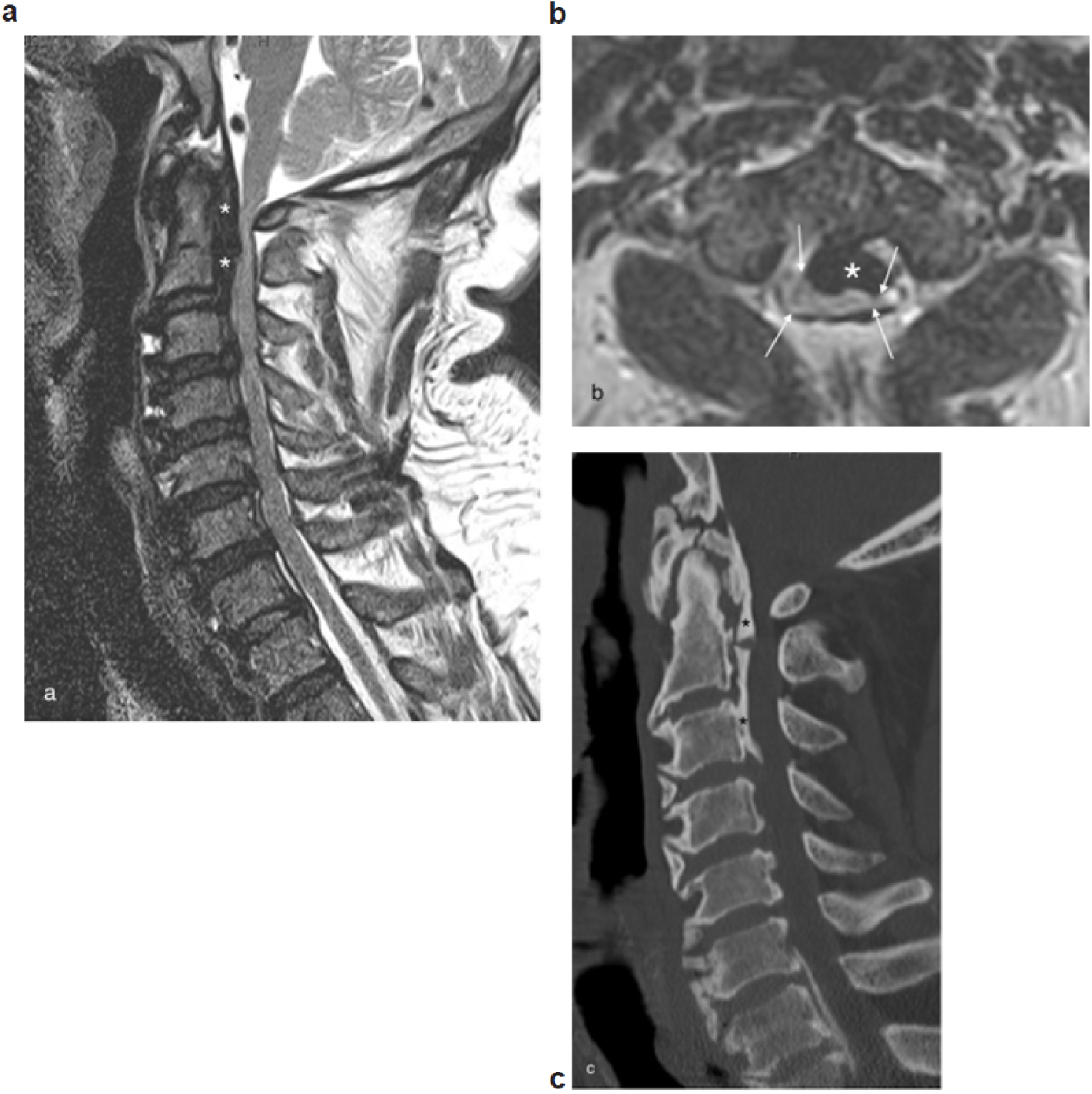
Magnetic resonance images. **a** Sagittal and axial **b** T2-weighted magnetic resonance images of the cervical spine demonstrate severe central stenosis primarily secondary to ossification of the posterior longitudinal ligament (OPLL) indicated by*. The spinal cord is severely compressed, left greater than right (white arrows). Sagittal reconstruction of computed tomography images **c** better demonstrates the ossification secondary to OPLL.

**Figure s4.**
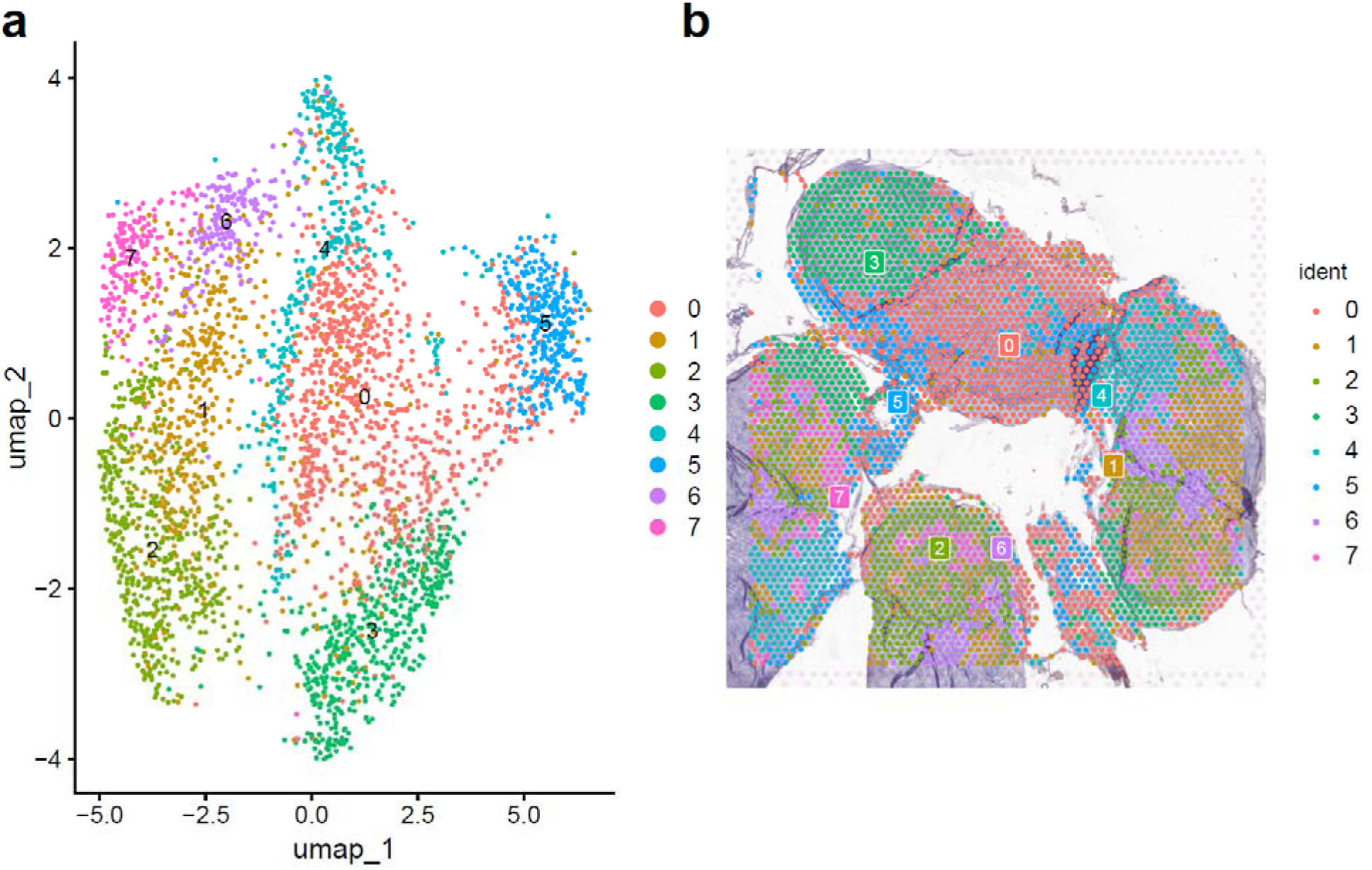
Spatial transcriptomic analysis of DRG tissue sections using Seurat in R. **a** UMAP visualization showing clustering based on the first 30 principal components. Each cluster corresponds to a distinct spatial location in the patient’s **<u>left</u>**side DRG tissue sections mounted on the Visium slide. **b** Spatial mapping of the identified clusters onto the tissue sections. A total of 8 clusters were identified, with Cluster 5 showing the highest expression of *IGHG4* (adjusted p < 0.0001, average log₂ fold change: 1.88). A log fold-change threshold of 0.2 was applied to identify differentially expressed genes.

**Figure s5.**
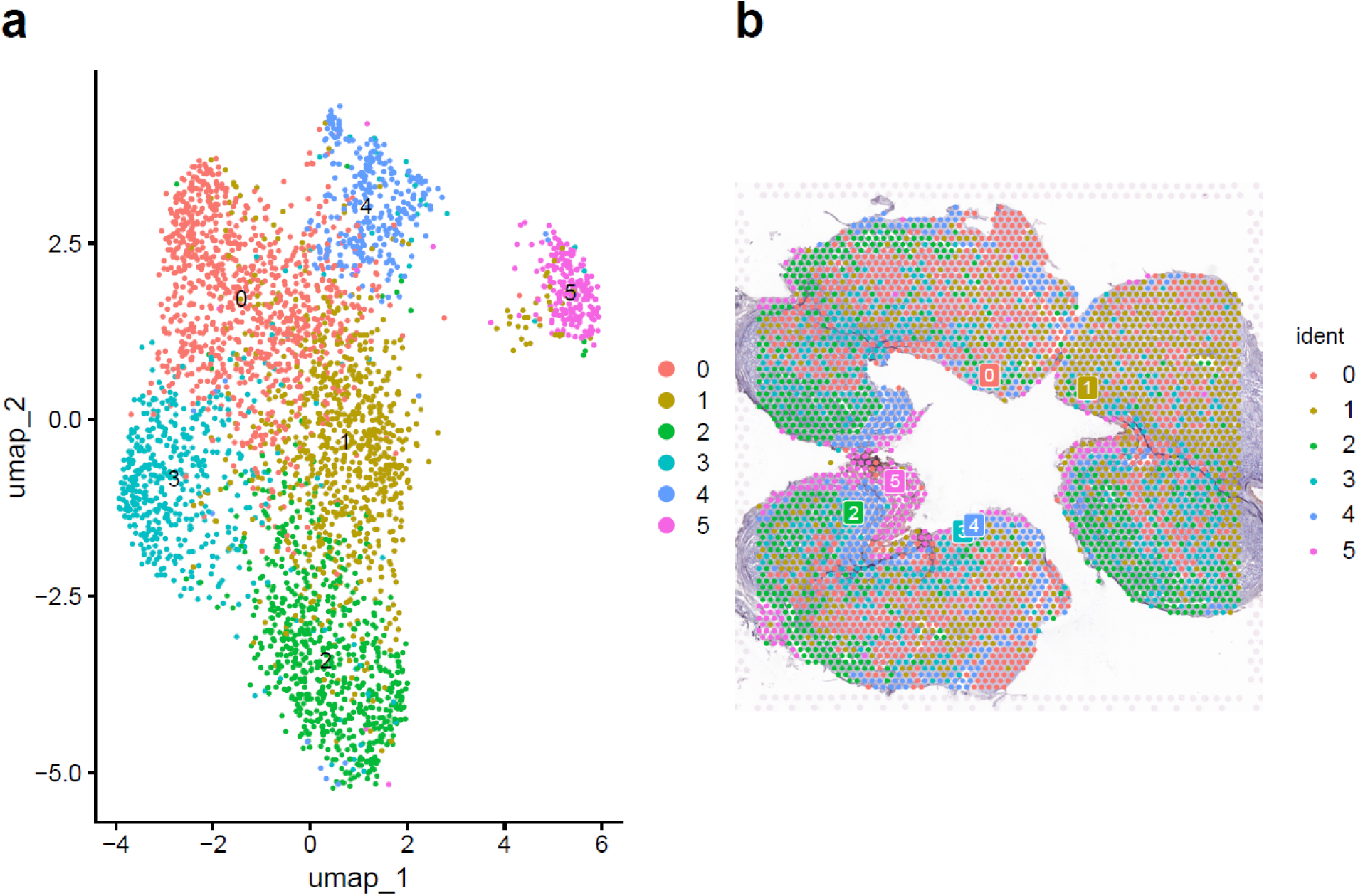
Spatial transcriptomic analysis of DRG tissue sections using Seurat in R. **a** UMAP visualization showing clustering based on the first 30 principal components. Each cluster corresponds to a distinct spatial location in the patient’s **<u>right</u>**side DRG tissue sections mounted on the Visium slide. **b** Spatial mapping of the identified clusters onto the tissue sections. A total of 8 clusters were identified, with Cluster 3 showing the highest expression of *IGHG4* (adjusted p < 0.0001, average log₂ fold change: 1.65). A log fold-change threshold of 0.2 was applied to identify differentially expressed genes.

**Figure s6.**
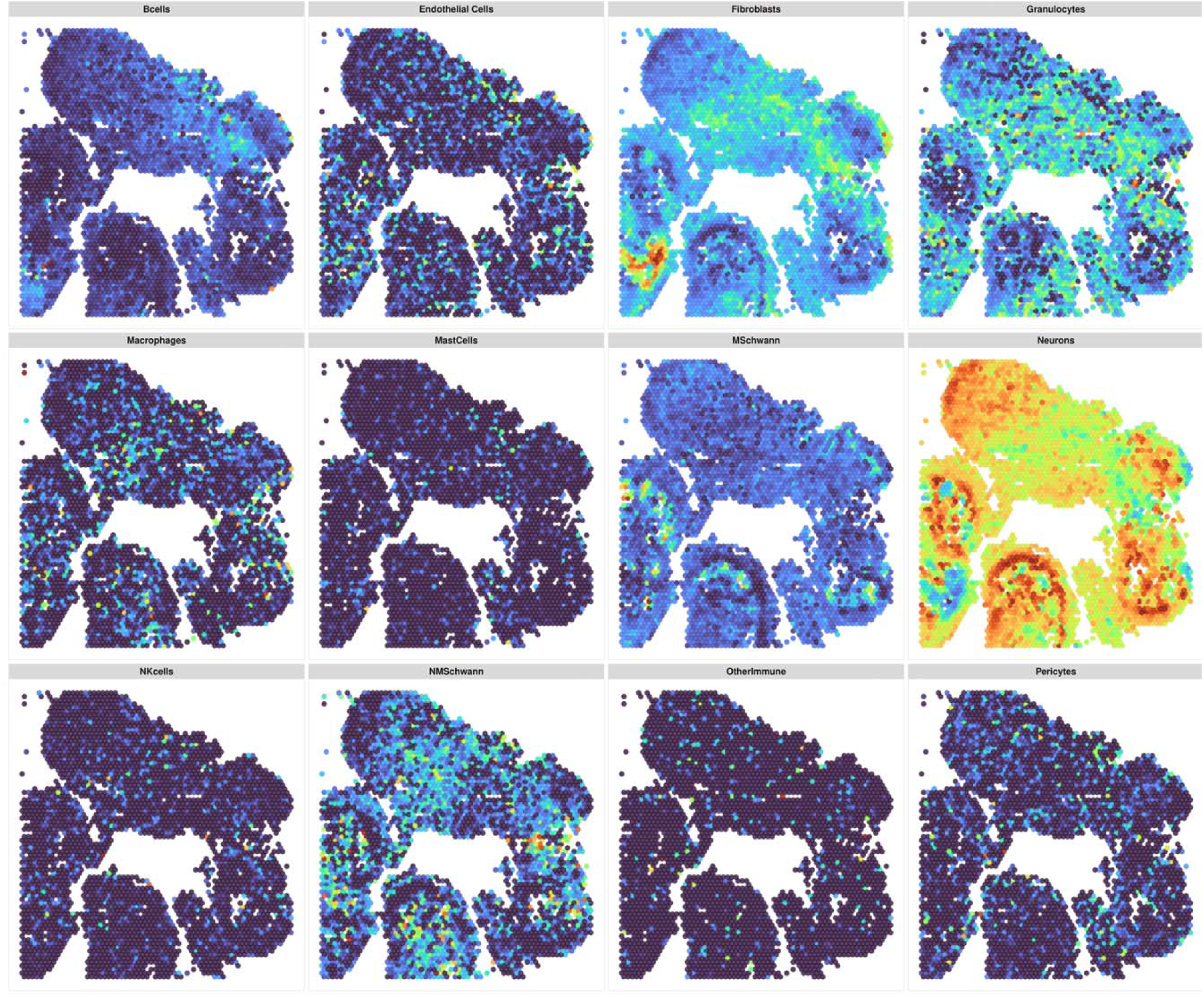
Computational cell type deconvolution using SONAR in R and MATLAB of the left C2 DRG. The figure shows 12 different cell type deconvolution patterns of the patient’s **<u>left</u>** side DRG. The proportion scale ranges from 0 (dark blue) to 1 (dark red).

**Figure s7.**
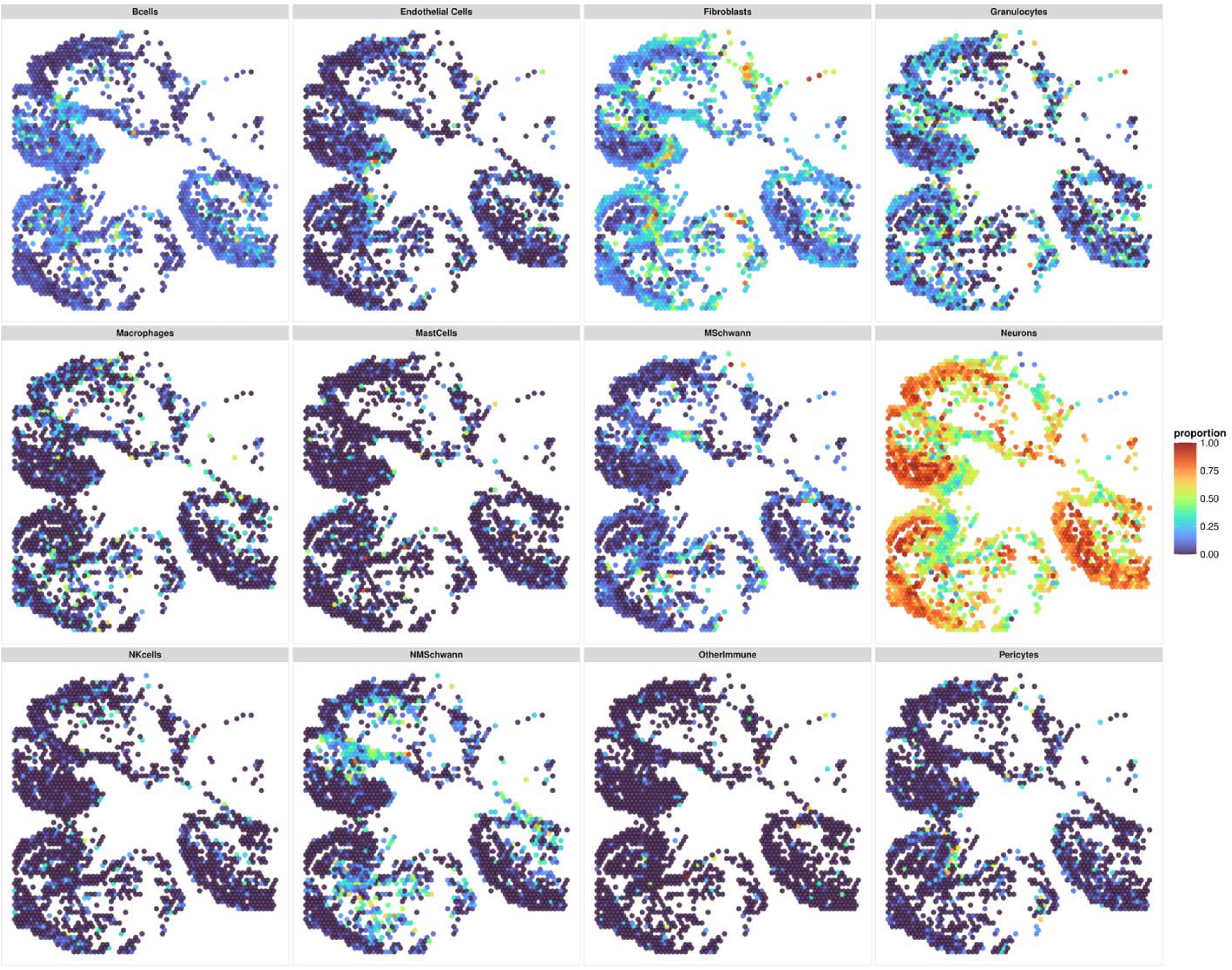
Computational cell type deconvolution using SONAR in R and MATLAB of the right C2 DRG. The figure shows 12 different cell type deconvolution patterns of the patient’s **<u>right</u>** side DRG. The proportion scale ranges from 0 (dark blue) to 1 (dark red). Missing barcodes are due to the exclusion of barcodes with low read counts.

**Figure s8.**
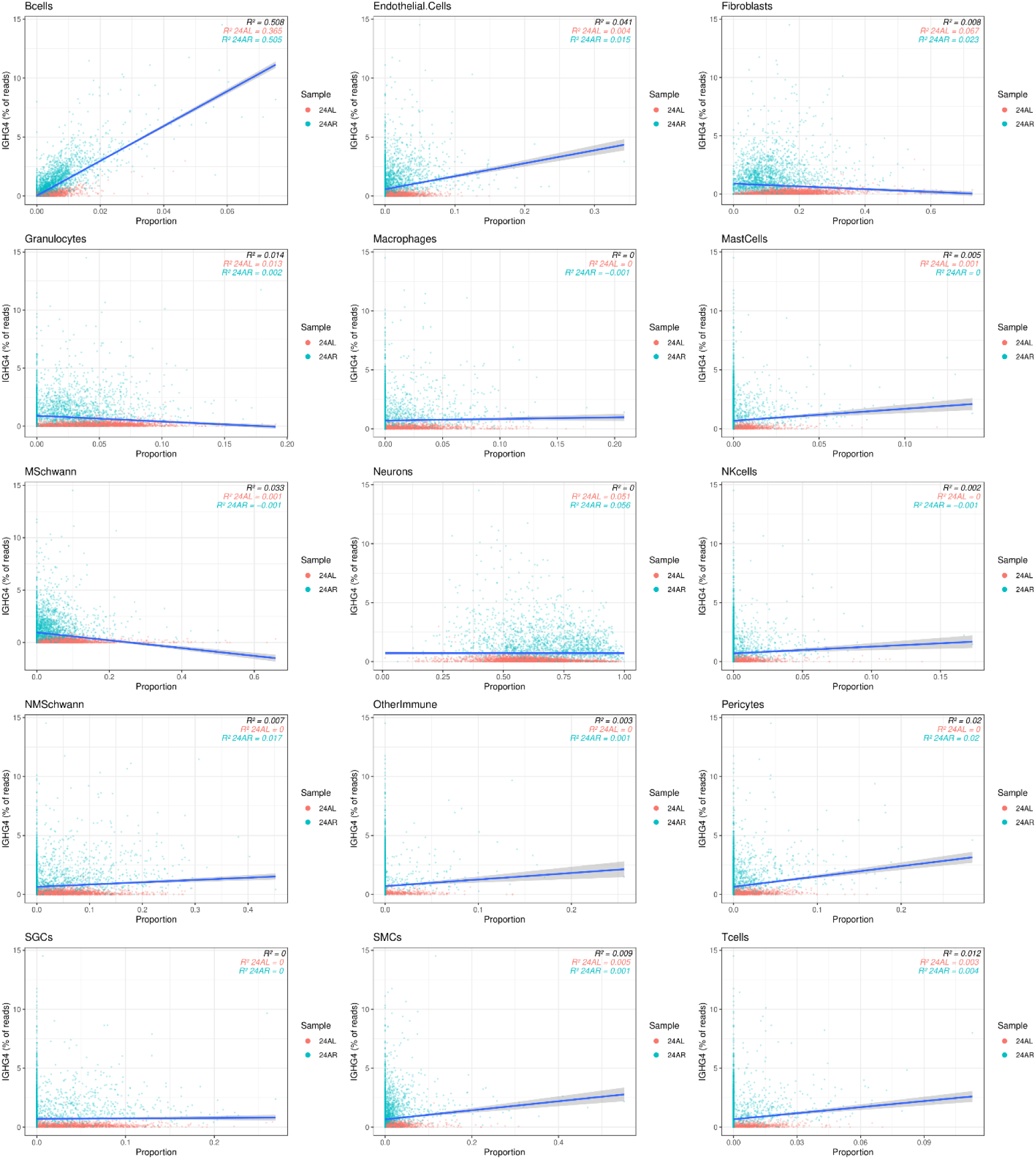
Correlation between cell type proportions and *IGHG4* expression. Each correlation plot displays the relationship between the proportion of one of 15 distinct cell types, and the percentage of *IGHG4* transcript reads on the left (red) and right (blue) sided DRGs of the patient. Each dot represents a unique spatial barcode.

**Figure s9.**
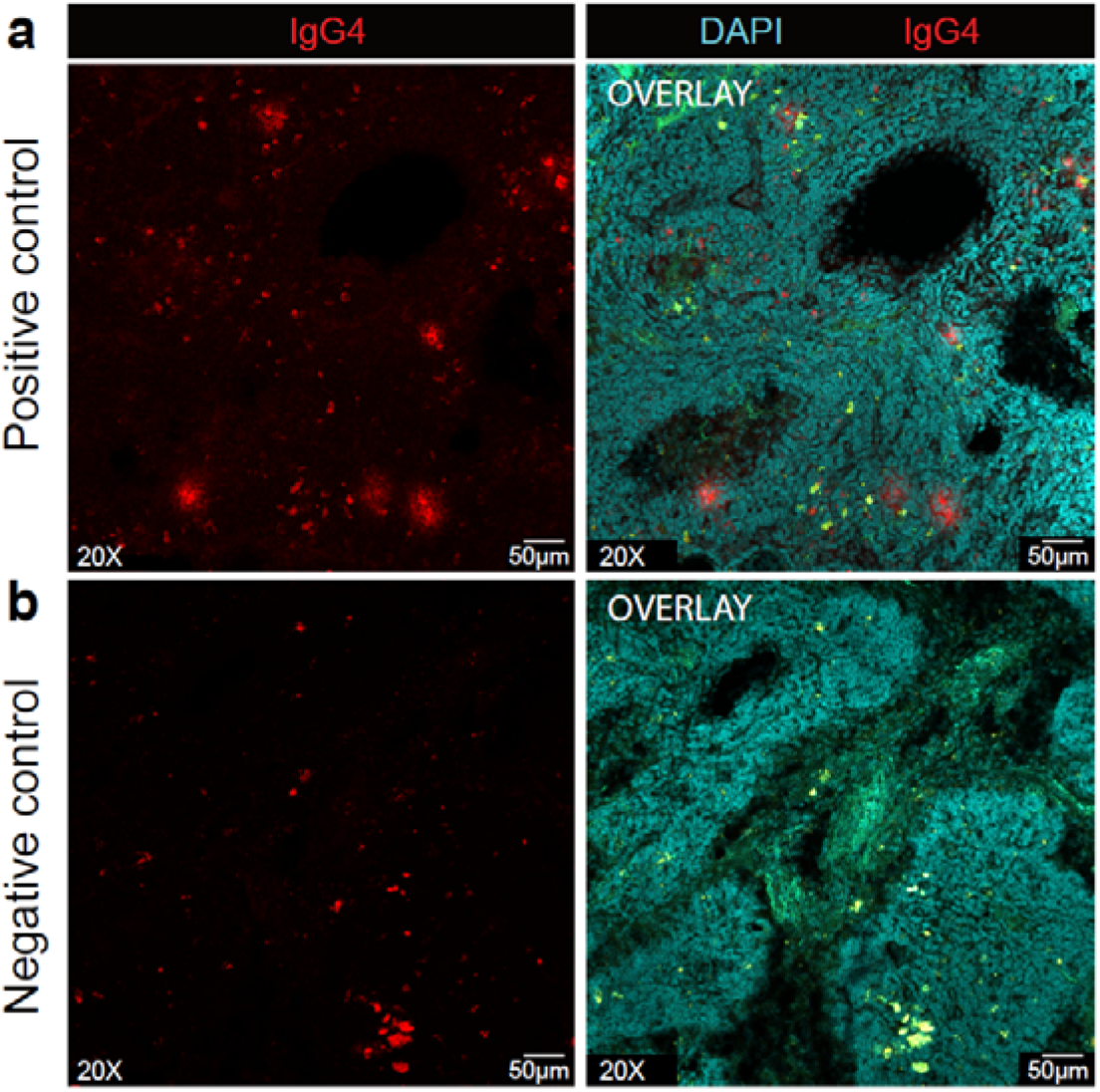
Optimization of anti-IgG4 antibody immunofluorescence staining in human lymph nodes. **a** Positive control staining with rabbit anti-IgG4 (1:1000, Abcam, ab109493) and DAPI (1:1000, Cayman Chemical Company, 14285). The left panel displays the IgG4 signal (red), while the right panel shows the overlay with DAPI staining (cyan). **b** Negative control staining without secondary antibodies. The left panel shows background signal, indicative of autofluorescence, while the right panel displays the DAPI staining for nuclear identification. The green signal in the left panel represents natural autofluorescence.

**Figure s10.**
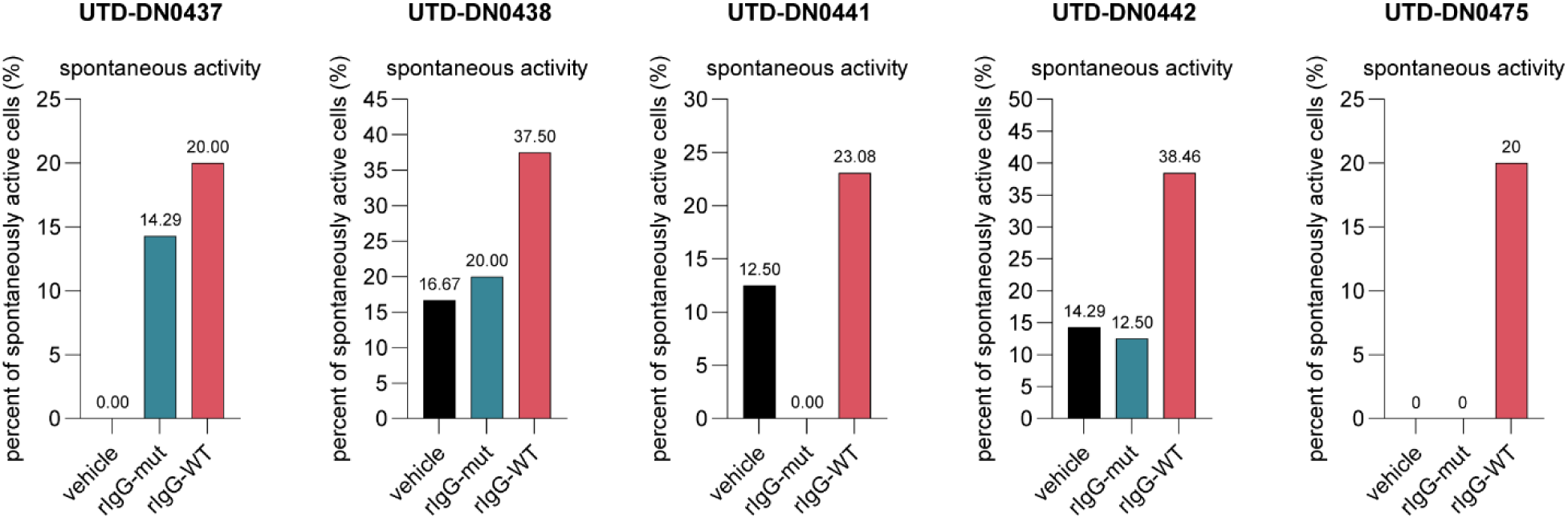
Electrophysiological recording of neuronal spontaneous activity per donor. Graph bars showing the percent of spontaneously active cells per donor across 3 groups: vehicle, rIgG-mut and rIgG-WT, where the rIgG-WT consistently has a higher percentage across donors in comparison to the other 2 groups.

**Supplemental Table 1.** Neurologic exam of case study patient.

| **Strength** | **Right** | **Left** |
| --- | --- | --- |
| Shoulder Abduction | 5/5 | 4/5 |
| Elbow Flexion | 5/5 | 4/5 |
| Elbow Extension | 5/5 | 4/5 |
| Wrist Extension | 5/5 | 4/5 |
| Grip | 5/5 | 4/5 |
| Finger Abduction | 5/5 | 4/5 |
| Hip Flexion | 5/5 | 5/5 |
| Knee extension | 5/5 | 5/5 |
| Dorsiflexion | 5/5 | 5/5 |
| Plantar flexion | 5/5 | 5/5 |
| **Light touch** | **Right** | **Left** |
| Lateral arm (C5) | Normal | Diminished |
| Lateral forearm/hand (C6) | Normal | Diminished |
| 3rd digit (C7) | Normal | Diminished |
| Medial hand (C8) | Normal | Diminished |
| Medial forearm (T1) | Normal | Diminished |
| Anteromedial thigh (L1, L2) | Normal | Diminished |
| Medial Knee (L3) | Normal | Diminished |
| Medial Malleolus/Calf (L4) | Normal | Diminished |
| Great toe/Lateral calf (L5) | Normal | Diminished |
| Lateral Foot (S1) | Normal | Diminished |
| **Reflexes** | **Right** | **Left** |
| Biceps (C5) | 3+/4 | 3+/4 |
| Brachioradialis (C6) | 3+/4 | 3+/4 |
| Triceps (C7) | 3+/4 | 3+/4 |
| Patellar (L4) | 3+/4 | 3+/4 |
| Achilles (S1) | 3+/4 | 3+/4 |
| Hoffman's | Absent | Positive |
| Clonus | Absent | 2 beats |
| **Additional findings** |  |  |
| Mental status | Alert and oriented x3 |  |
| Motor tone | normal |  |
| Atrophy/Fasciculations | absent |  |
| Station and gait | Antalgic gait, not wide based |  |

**Supplemental Table 2.** Further Clinical Characteristics.

| Measure | Score |
| --- | --- |
| Neck Disability Index (range 0-24) | 12 |
| Depressive symptoms (PHQ-2) (range 0-6) | 4 |
| Anxiety symptoms (GAD-2) (range 0-6) | 1 |
| Catastrophizing (PCS-4) (range 0-16) | 10 |
| **Current medications for pain:** |  |
| - Oral acetaminophen 325 mg 2 every 6 hours<br>- Oral gabapentin 100 mg every 8 hours |  |

PHQ-2: Two item Patient Health Questionnaire.<sup>1</sup>
GAD-2: Two-item Generalized Anxiety Disorder.<sup>2</sup>
PCS-4: Four-item Pain Catastrophizing Scale.<sup>3</sup>

**Supplemental Table 3.** Gene panel used for identification of B cells deconvolution with SONAR.

| Gene | cluster |
| --- | --- |
| TNFRSF13B | B cells |
| MS4A1 | B cells |
| BLK | B cells |
| DEPRECATED-ENSG00000211592 | B cells |
| IGKC | B cells |
| CD79A | B cells |
| LY9 | B cells |
| BCL11A | B cells |
| NIBAN3 | B cells |
| HSH2D | B cells |
| TNFRSF13C | B cells |
| LTB | B cells |
| P2RY10 | B cells |
| CD52 | B cells |
| C16orf54 | B cells |
| SYTL1 | B cells |
| IKZF3 | B cells |
| BANK1 | B cells |
| PARP15 | B cells |
| ACAP1 | B cells |
| CYTIP | B cells |
| CCDC88C | B cells |
| RHOH | B cells |
| TBC1D10C | B cells |
| ATP2A3 | B cells |
| WDFY4 | B cells |
| SEL1L3 | B cells |
| IL2RG | B cells |
| CD37 | B cells |
| PTPN22 | B cells |

**Supplemental Table 4.** Antibodies used for immunohistochemical experiments.

| Antibody | Dilution | Vendor | Catalog number |
| --- | --- | --- | --- |
| Anti-IgG4 EP4420 | 1:1000 | Abcam | ab109493 |
| Anti-Peripherin | 1:1000 | EnCor Biotechnology | CPCA-Peri |
| Anti-CD64 | 1:250 | Invitrogen | 14-0649-82 |
| goat anti-rabbit Alexa<br>Fluor 555 | 1:2000 | ThermoFisher<br>Scientific | A-21428 |
| goat anti-chicken<br>Alexa Fluor 488 | 1:2000 | ThermoFisher<br>Scientific | A-11039 |
| goat anti-mouse<br>Alexa Fluor 555 | 1:2000 | Invitrogen | A-21137 |

**Supplemental Table 5.**
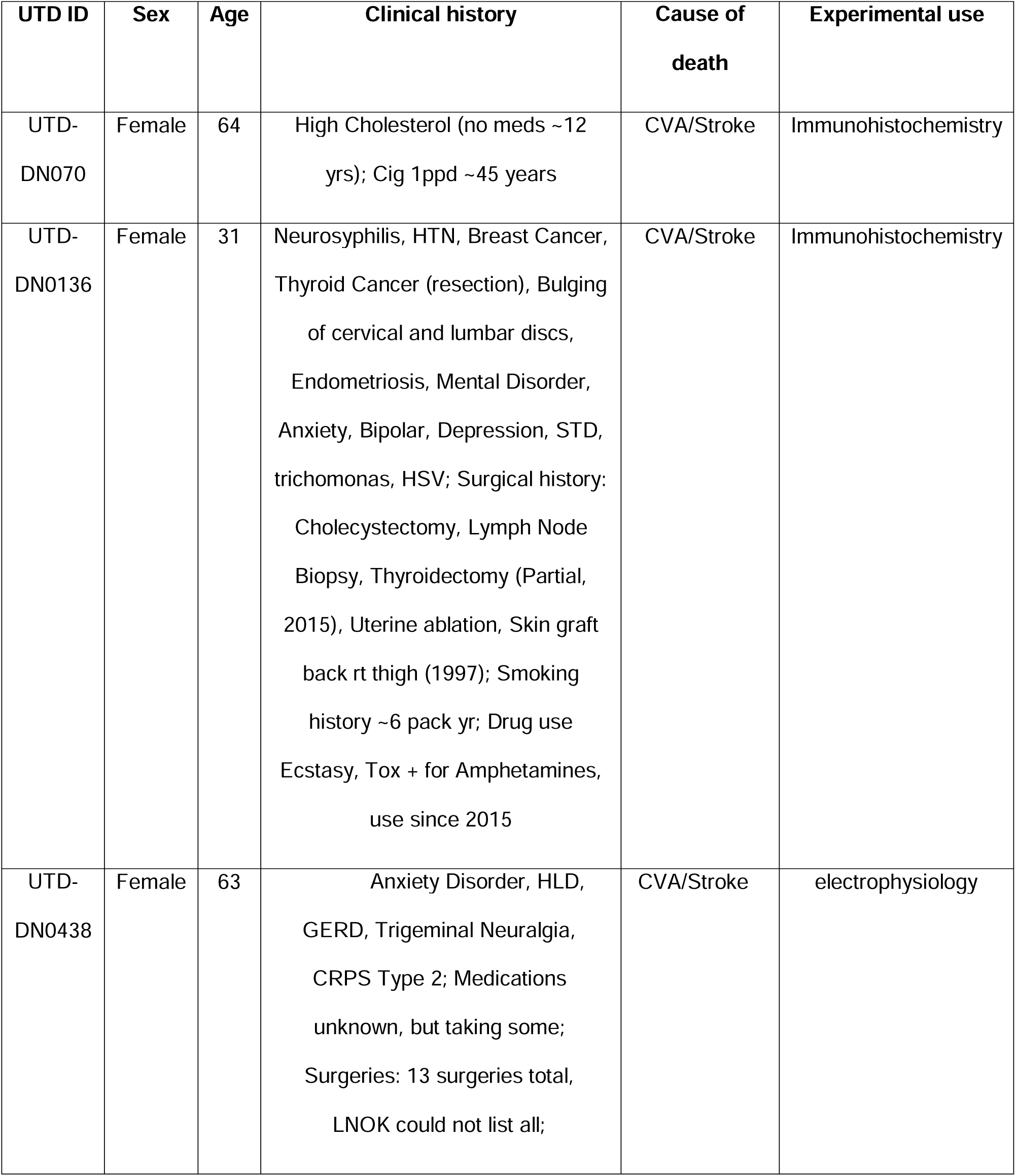
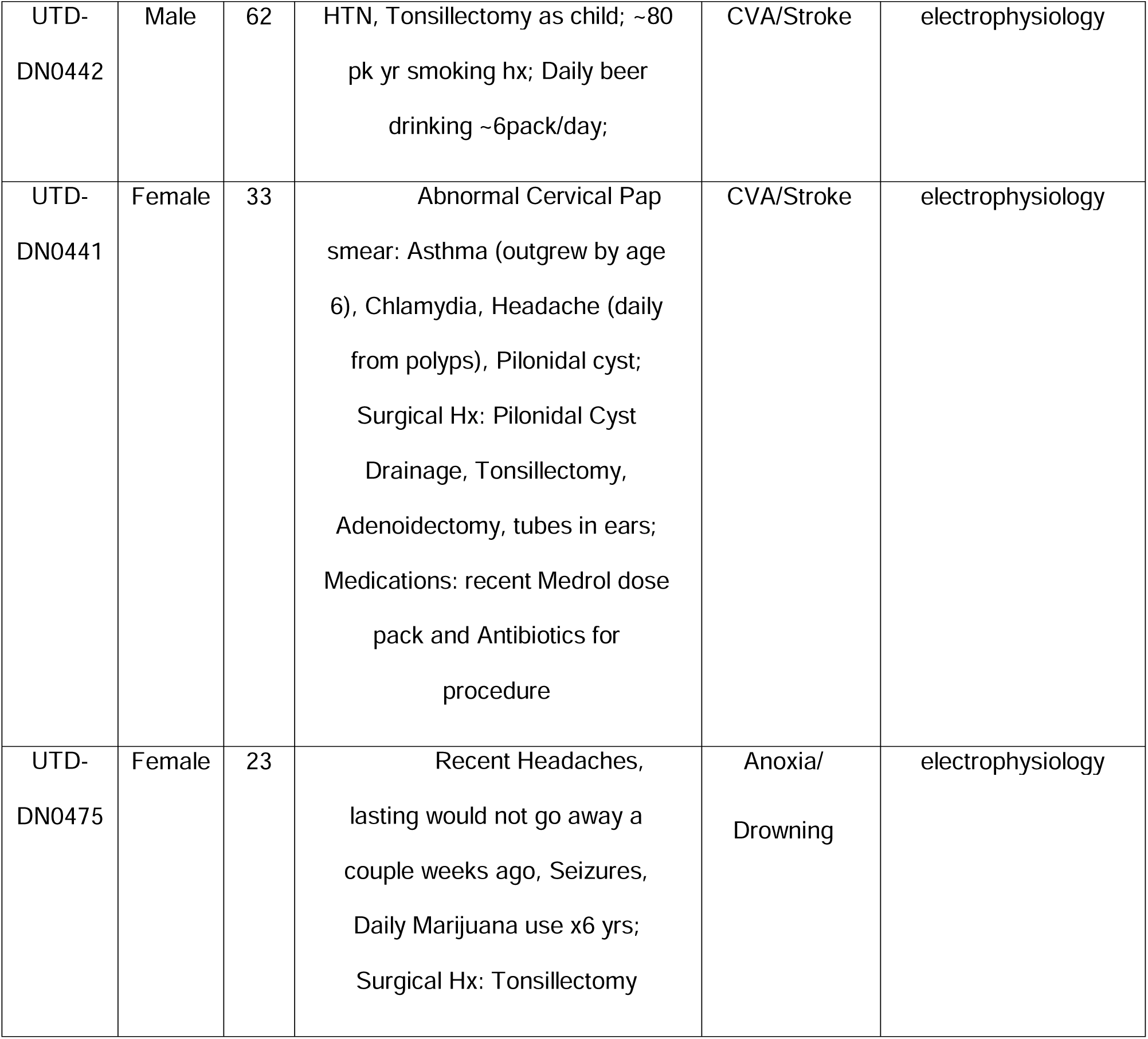
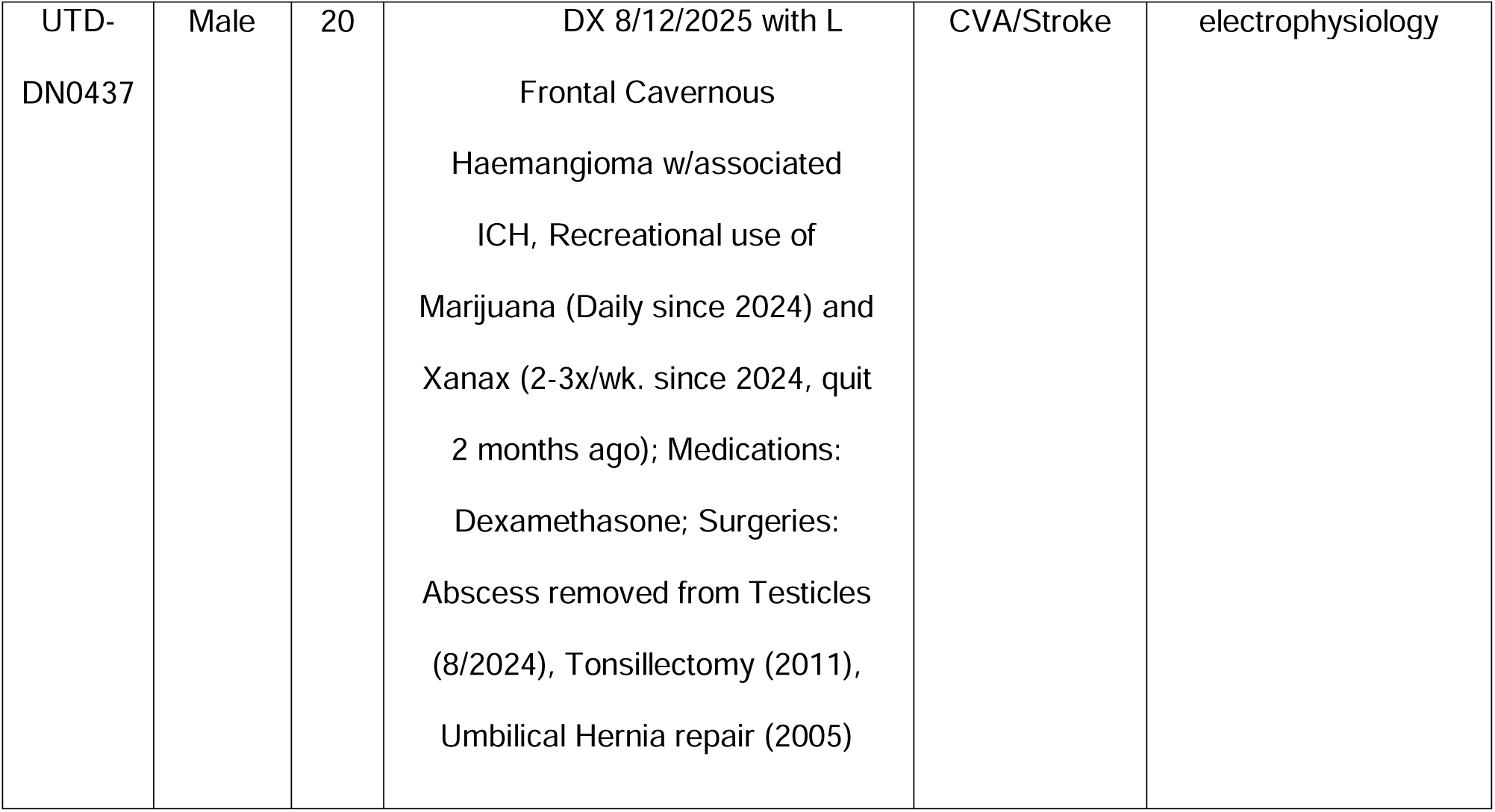
Donors’ demographics for immunohistochemical and electrophysiological experiments.

## Supplementary methods

### Participants and setting

Patients were identified from emergency care settings and clinic visits. We screened patients who were older than 18 years and undergoing arthrodesis that included the C1-C2 segment. Patients were identified in a two-pronged manner, whereby the research staff were notified by the surgeon of scheduled procedures and received daily electronic reports accessing the feeds of surgery scheduling data, using the UW Medicine Enterprise Data Warehouse. Exclusion criteria were cognitive deficits and language comprehension that would make the patient unable to consent. Potentially eligible outpatients were contacted via phone call to set up a time to meet in person. Inpatients were met in their hospital room if they indicated interest in participating in the study. The research staff then informed the participants of the study and obtained informed consent before starting the study procedures.

### Clinical characteristics

Age, sex, body-mass index (BMI), race, and ethnicity were recorded within two weeks of the surgery from the electronic health records (EHR) when available or were otherwise confirmed by patient interview.

Patients were asked to rate their average neck/occipital pain separately for the left and right side during the last 24 hours using a numerical rating scale (NRS), with 0 = no pain and 10 = worst imaginable pain^1^. They indicated the area of pain in a body chart, as well as the duration of pain, which was recorded in days. Participants were categorized as having acute (lasting <3 months) or chronic pain (≥3 months), in keeping with the definitions of the International Association for the Study of Pain (IASP)^2^.

Neck-related function was assessed using the 5-item version of the Neck Disability Index (NDI-5)^3^. Depressive and anxiety symptoms were assessed using the two-item Patient Health Questionnaire (PHQ-2)^4^ and the two-item Generalized Anxiety Disorder (GAD-2)^5^. Catastrophizing was assessed using the four-item Pain Catastrophizing Scale (PCS-4)^6^.

Current medications used for pain treatment and non-pharmacological treatments related to pain and to mental health comorbidities were abstracted from the EHR and confirmed by patient interview. Additionally, participants were specifically asked about any opioid, anticonvulsant, or antidepressant use in the preceding 6 months.

Pre-operative imaging consisted of a cervical spine magnetic resonance imaging (MRI) examination obtained on a Hitachi open 1.2 tesla system using the following sequences: sagittal and axial T1- and T2-weighted, sagittal short tau inversion recovery (STIR), axial phase balanced Steady State Acquisition Rewound Gradient Echo (PBSG).

Computed tomography (CT) was acquired on a Siemens Emotion 16 scanner with 1mm thick axial sections reconstructed in both soft tissue and bone algorithms as well as 2mm thick sagittal and coronal multiplanar reformations.

The patient with high levels of *IGHG4* in the C2 DRG was male, Pacific Islander, and non-Hispanic. He underwent a posterior C1-T2 fusion and bilateral C1-7 laminectomy, with resection of the C2 DRG bilaterally.

The patient started experiencing left neck and arm pain 2 years before surgery and then reported spread of pain to the right side of his neck and both extremities. Prior to surgery, he rated his average pain intensity in the past 24 hours as 8.0 on the left side of the neck and 6.5 on the right side, and described the pain as radiating bilaterally to the shoulders, arms, fingers, and legs. Additionally, he experienced numbness and weakness bilaterally in both upper and lower extremities, difficulties in holding on to objects, handwriting, and walking, associated with balance difficulties, prompting him to use a cane for ambulation assistance, but had no history of falls or incontinence.

The neurologic examination showed sensory and motor impairments of the 4 extremities, more marked on the left side (**Supplemental Table 1**). A single neuroradiologist (J.G.J.),blinded to the patient’s clinical characteristics other than knowledge that he had bilateral neck pain, evaluated the patient’s imaging. The MRI and CT showed severe central stenosis with severe cord compression at C1-2, C2-3, and C3-4, and moderate stenosis at C4-5, C5-6, and C6-7, secondary to ossification of the posterior longitudinal ligament that involved all cervical levels (**Fig. 1**). The spinal cord compression and central stenosis were generally more severe on the left side. The cervical and thoracic spine also showed extensive diffuse idiopathic skeletal hyperostosis, including ossification of the ligamentum flavum. Further clinical characteristics are reported in **Supplemental Table 2**.

Since surgery, the patient’s pain and neurological deficits have not improved, as documented in follow-up visits up to three years after surgery.

### C1-C2 Arthrodesis patients: recruitment and tissue recovery

Patients undergoing C1-C2 arthrodesis for treating neck pain at the University of Washington were recruited for a study approved by the University of Washington Internal Review Board (study 10916). Details on the study procedures are provided in **Supplemental File 1**. Bilateral C2 DRGs from patients were harvested during neurectomies and fresh-frozen with pulverized dry ice^7^. The tissues were shipped to the University of Texas at Dallas (UTD) for further RNA sequencing and processing.

### Immunocytochemistry

We followed a modified version of the immunohistochemistry protocol described by Yousuf et al.,^8^. Human lymph nodes sections were added as positive control. Cryosections of 10 µm thickness were prepared at −20°C from DRGs and lymph nodes previously embedded in OCT and mounted on charged SuperFrost slides. The sections were fixed with 10% buffered formalin (Thermo Fisher Scientific, 23245684) for 10 minutes, followed by a series of four dehydration steps using 50%, 70%, and two rounds of 100% ethanol (200 proof, Thermo Fisher Scientific, 07678005) for 5 minutes each.

Next, the tissues were blocked for 1 hour in a blocking buffer composed of 10% heat-inactivated donor goat serum (NGS) (R&D Systems, S13150H) in PBS-TX (0.1% Triton X-100 (Sigma-Aldrich, T8787) in 1X phosphate-buffered saline (PBS) (Thermo Fisher Scientific, BP39920). All antibodies were prepared in an antibody stock solution containing 2% bovine serum albumin (BSA) (EMD Millipore, 126609), and 2% NGS in PBS-TX. Primary antibodies (**Sup. Table 4**) were applied to the slides and incubated overnight at 4°C. The slides containing lymph node sections were incubated exclusively with Anti-IgG4. The slides were then washed twice with PBS-Tween (0.1% Tween-20 (Fisher Scientific, BP337) in 1X PBS) (Thermo Fisher Scientific, BP337500) and once with 1X PBS for 5 minutes per wash.

For secondary antibody (**Sup. Table 4**) incubation, the tissues were treated for 1 hour at room temperature in a dark humidity chamber. After incubation, the slides were washed again with two rounds of PBS-Tween and one round of 1X PBS.

Nuclear staining was performed using DAPI (1:1000, Cayman Chemical Company, 14285) in PBS-TX for 5 minutes, followed by a final 5-minute wash in 1X PBS. The slides were then mounted with ProLong Gold Antifade Mountant (Thermo Fisher Scientific, P36930) and 1.5 mm coverslips (Harvard Apparatus, 640707). Imaging was conducted using an FV4000 confocal laser scanning microscope (Evident Scientific) at 20x magnification. For display purposes, the raw image files were brightened and adjusted for contrast using Olympus CellSens software (v1.18).

### Electrophysiology

In current clamp, a 90 second recording with no current injected was taken to assess spontaneous activity. If a cell fired spontaneously, no further recordings were taken. Rheobase was then measured using a series of 20-millisecond current steps increasing current by 10 pA until the cell fired the first action potential. Next, a series of 1-second ramp current injections was applied to the cell, starting with a ramp from 0-100 pA and increasing the ramp ending current by 200 pA up to maximum of 2900 pA. Finally, a series of 500-millisecond step current injections starting with 0 pA and increasing by 50 pA up to a maximum of 2100 pA was injected. Data were analyzed by the experimenter using Clampfit 10.4 software.

Spontaneous activity was assessed in current clamp mode for the first 90 seconds without any stimulus. Proportion of spontaneous cells was defined as the number of spontaneously firing neurons per the total number of cells that were recorded from. Spontaneously firing cells were not used for subsequent analysis. Current ramps, rheobase, threshold, amplitude, half-width, afterhyperpolarization, rising slope, and falling slope were calculated as previously reported^9^.

## Notes

### Competing Interest Statement

The authors have declared no competing interest.

### Author Declarations

The study was approved by the University of Washington Internal Review Board (study 10916).

### Summary of Updates

We have expanded the study by adding in-vitro validation and mouse tranffer study.

